# Evaluation of the effects of vaccination regimes on the transmission dynamics of COVID-19 pandemic

**DOI:** 10.1101/2022.01.22.22269569

**Authors:** Ichiro Nakamoto

## Abstract

The COVID-19 pandemic has yet to be eliminated globally despite the advancement of immunization programs. Evaluation of the effects of the vaccination regimes of COVID-19 is critical for understanding the potential capacity of countermeasures and informing subsequent prioritization strategies of responses. Research and observational data provide broad support regarding the importance of effective vaccines, in contrast, debates remain on the timing and priority of booster vaccination under the assumption of resource constraint. This study aims to evaluate the effect of vaccination regimes on the trajectory of the COVID-19 pandemic from the medium-term perspective. We employ a mathematical model to infer critical epidemiological characteristics associated with COVID-19, thereafter perform simulation on the transmission dynamics of the epidemic up to 3 years. The outcomes imply that in the absence of severe variants of the pathogen, administration of booster vaccination curtails the peak size of total cases and share of severe infections at later waves. Nevertheless, it can be better off by prioritizing the primary doses to unvaccinated individuals when vaccine shortage is challenged. The effects of priority categories are consistent across a broad range of profiles. Increasing the rollout capacity (i.e., administration rate) of doses can render the reproduction number lower than one and hence contain the transmission of pandemic ultimately controlling for other factors. The timing of rollout of primary doses is pivotal in reducing the magnitude of transmission saturation. It is of importance to prioritize the administration of primary vaccination series to vulnerable individuals efficiently and thereafter increment of administration capacity when the supply of vaccine increases over time to scale down the size of an epidemic.

---

By the end of 2021, nearly 0.3 billion cases of COVID-19 infections worldwide were officially identified, of which over 5 million deaths were confirmed in more than 200 countries and areas ^1^. The likelihood of containing the pandemic is low until the global availability of effective vaccines for the general population ^2^. Equitable and timely vaccination is supposed to play critical roles in controlling the spread of the SARS-CoV-2 epidemic ^3^. The Emergency Use Authorization of vaccines has expedited the progress of large-scale vaccine deployment ^4,5^. And a total of billions of doses have been administered as of January 2022 according to the report by WHO ^1,6^. To date, nearly 170 countries or equivalently roughly five percent of the global population have reported official records of vaccination. Many countries have unleashed vaccination campaigns to provide partial immunity to the population, in contrast, more than one-half of the population in low-income countries remain unvaccinated (over 160 doses per 100 individuals for high- and medium-income countries versus below 20 doses per 100 individuals for low-income countries as of January 2022) ^7,8^. This reflects the remarkable challenge of universal accessibility and global allocation of COVID-19 vaccines ^2,9^.

Vaccines can protect recipients from infections by promoting the formation of immunity ^10,11^. Trials aiming to identify the efficacy and safety of vaccines have been extensively conducted. Studies so far found that primary vaccination series of COVID-19 (presently one or two doses in most cases) could achieve a variety level of protection between 60% and 95% with acceptable safety profile ^1,10,11^. Vaccines such as a two-dose regimen of BNT162b2 were capable of conferring efficacy over a period of weeks ^12^. Mass vaccination of the first doses of vaccines including Pfizer/BioNTech, Moderna and Oxford/AstraZeneca was associated with sizable reductions in hospital admissions ^10,11,13^. The primary two-dose vaccines by the former administered 3 weeks apart were documented with decently toleration. When treated with the first dose, the establishment of immunity protection was generally confirmed ^14^. For other vaccines such as Ad26.COV2.S, tests revealed that a single dose was effective against hospitalization and mortality ^15^. Other follow-up data of participants in a double-blinded trial reinforced the evidence of primary effectiveness as well ^16^.

On the other hand, vaccine immunity wanes over time naturally ^13^. Precise and systematic quantifying the dynamics are challenging because the degree of waning and remaining protection potentially hinge on complex observed and unobserved factors and could vary from person to person and diversify from vaccine to vaccine. Some studies have made substantial progress in this direction ^17^. Recent investigation indicated that vaccine effectiveness against severe COVID-19 decreased by almost 10% over an interval of 6 months ^18,19,20,21,22^. In older adults, protection against symptomatic disease decreased more over the same period. Similar waning protection of hospital admissions and mortality was also delineated ^19,20,21^. As a countermeasure, data revealed that participants who received a booster vaccine >5 months after the second dose had almost one-fold lower mortality due to COVID-19 ^18,21,22^. Other observational data identified that under the three-dose schedule, seroconversion rates of neutralizing protection two weeks after the third dose could reach a level of 97% ^19,20,23^. The booster vaccination played a sizable role in mitigating the burden of the past outbreak of Haemophilus influenza ^24^.

The topic of booster vaccination is controversial. For COVID-19, debates hover over the necessity, priority, and timing of booster dose for individuals who previously received the primary vaccination ^25,26^. Another public concern is the mechanism of the rates of administration and their interaction on affecting the transmission dynamics of pandemic essentially ^27^.

We evaluate the medium-term effects of booster vaccination regimes on the transmission dynamics of COVID-19 coupled with the traits of the vaccines, characteristics of the pathogen, and rate of vaccination. We employ a mathematical model to investigate (1) whether booster vaccination could reduce the ratio of cases and share of severe infections in the absence of severe variants; (2) the priority of vaccination strategy under dose supply constraints (i.e., efficiency-upgraded primary vaccination versus booster dose); and (3) whether vaccination rates and their interaction impact the transmission dynamics and fundamentally diminish the COVID-19 pandemic. To account for a variety of scenarios, we additionally assume that (1) the population is divided into two categories, the first category includes susceptible individuals who are vaccinated with routine primary doses and thereafter potentially the provision of booster vaccination, and the second category consists of unvaccinated individuals; (2) vaccine supply constraint is introduced at the point of strategy decision regarding the priority of booster vaccination or efficiency-enhanced primary vaccination; (3) the first dose of primary series confers partial and weaker immunity protection versus subsequent doses; and (4) the immunity conferred by doses wanes over time (Fig. 1).

**Fig 1.**
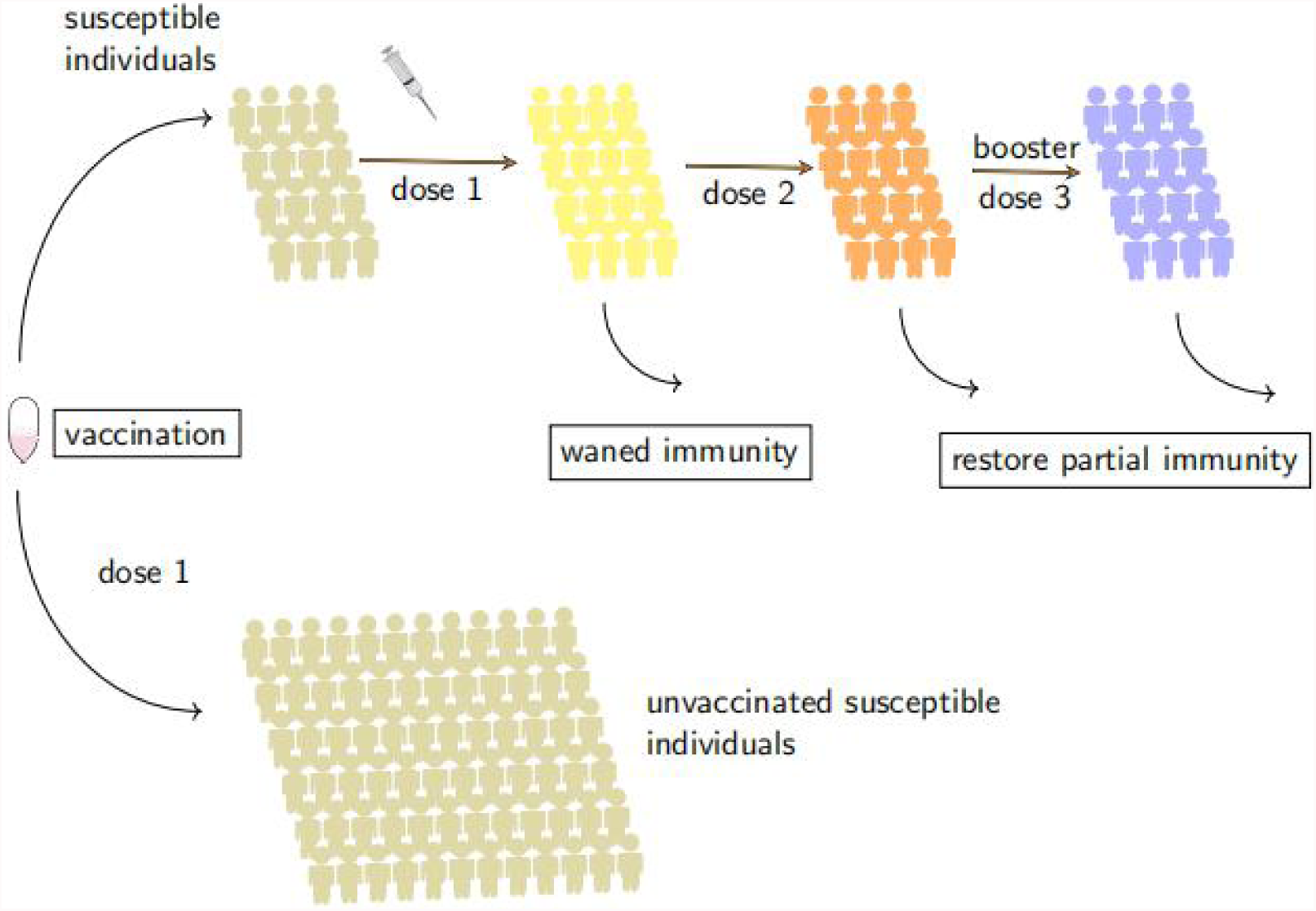
Vaccine doses regime flowchart. Individuals are divided into two groups, individuals of the first group are vaccinated with primary vaccine series and thereafter potentially booster dose, individuals of the second group are unvaccinated due to a variety of reasons including vaccine hesitancy (e g., concern of vaccines’ safety, concern of vaccines’ effectiveness, changed vaccine guidance, public confidence), supply inadequacy, supply imbalance or deployment inefficiency. Booster dose partially restores the immunity. The immunity of primary vaccination and booster dose wanes over time.

We show that the administration of booster dose reduces the ratio of total incidences and severe infections of COVID-19 in the absence of severe variants. However, increasing the availability and rollout rate of the primary doses to more vulnerable and unvaccinated populations need to be taken with priority over the booster dose strategy when supply inadequacy or imbalance is at work. A highly effective transmission-blocking primary-dose vaccine prioritized to the unprotected individuals reduces the peak size of incidences and severe disease outcomes. The vaccination prioritization strategy findings are broadly consistent across a variety of scenarios accounting for transmission rates, rollout speeds, immunity protection and dose spacing. The principal framework provides meaningful insights to compare the asymptotic impacts of prioritization strategies across various settings.

## Results

### Implementation of booster vaccination can reduce the incidence of total cases and severe infections

We simulate the effect of booster vaccination on the transmission dynamics of COVID-19 under a variety of scenarios (Fig. 2a-2h) and sensitivity analysis is tested. The rollout initiation of dose 1 is assumed to start at week 45, nearly one year after the onset of the pandemic outbreak. The efficacy of vaccines has been identified in trials albeit the presence of heterogeneity in levels ^12,13,15,20,23^. Some studies recommend 3 or 4 weeks of spacing between dose 1 and dose 2 ^20,5,28^. Others document that receipt of different doses or varying dose spacing can induce decent protection as well ^17,10,14,20,26^. We evaluate a diversified range of dose 1&2 spacing where the distribution is of 3(Fig. 2c-2f,2h), 8(Fig. 2a-2b), and 12(Fig. 2g) weeks respectively. Prior work suggests that a booster dose of mRNA vaccine >5 months apart is effective in partially restoring waning immunity and protecting individuals against severe COVID-19 outcomes in comparison with the primary two-dose strategy ^29^. We appraise the scenario where the spacing of doses 2&3 is 24 weeks (Fig. 2a,2b,2c,2e,2g), and extend this to 32(Fig. 2d) and 48 weeks(Fig. 2f). To assess whether the earlier deployment of dose 3 exerts a differentiated effect, we decrease the length of 2&3 spacing to 8 weeks thereafter. The immunity of dose 1 declines over time and vaccinated individuals are exposed to infection risk consequently, the length of which is presumed as 6.5 weeks, and for doses 2 and 3 the hypothesized value varies from 26 to 52 weeks respectively. We delay the rollout initiation of dose 1 for 2 weeks to test the sensitivity of vaccination timing(Fig. 2e). Two differential administration rates of dose 3 (0.02 versus 0.05 per week) are utilized to mimic expeditious and speed-compromised administration contexts respectively. The parameters utilized in the simulations are outlined in detail (see Supplementary Table S1).

**Fig 2.**
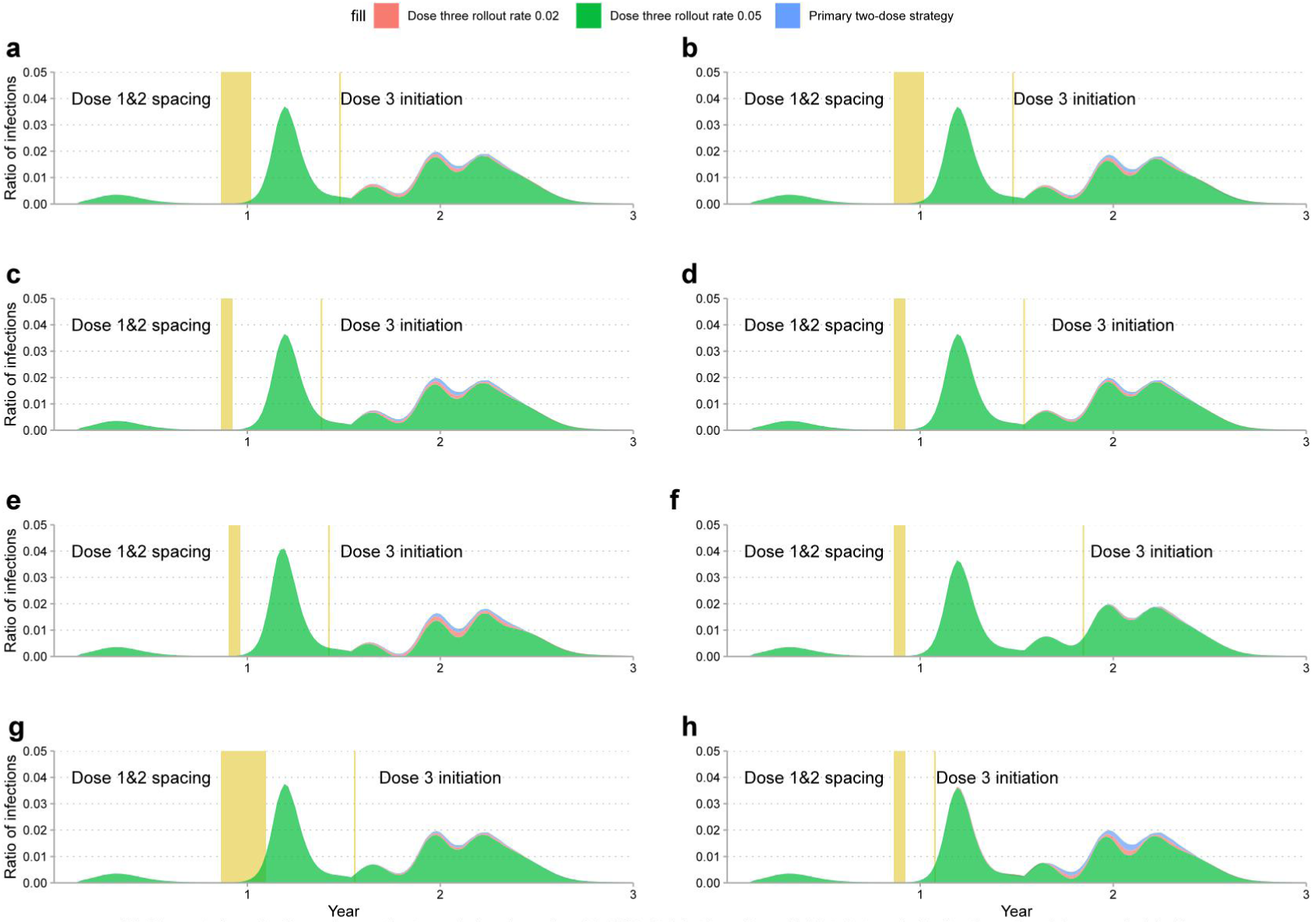
Impact of vaccination regime on the transmission dynamics of COVID-19 infections. Dose 1 initiated at week 45 after the outbreak in scenario (a)∼(d) and (f)∼(h). Initiation of dose 1 in scenario (e) delayed to week 47.The immunity conferred by dose 1 wanes to susceptibility after 6.5 weeks in all scenarios, (a) Dose 1&2 spacing 8 weeks and 2&3 spacing 24 weeks, immunity conferred by dose 2&3 wanes to susceptibility after 26 weeks; (b) Dose 1&2 spacing 8 weeks and 2&3 spacing 24 weeks, immunity conferred by dose 2&3 wanes to susceptibility after 52 and 26 weeks respectively; (c) Dose 1&2 spacing 3 weeks and 2&3 spacing 24 weeks, immunity conferred by dose 2&3 wanes to susceptibility after 26 weeks; (d) Dose 1&2 spacing 3 weeks and 2&3 spacing 32 weeks, immunity conferred by dose 2&3 wanes to susceptibility after 26 weeks; (e) Dose 1&2 spacing 3 weeks and 2&3 spacing 24 weeks, immunity conferred by dose 2&3 wanes to susceptibility after 26 weeks; (f) Dose 1&2 spacing 3 weeks and 2&3 spacing 48 weeks, immunity conferred by dose 2&3 wanes to susceptibility after 26 weeks; (g) Dose 1&2 spacing 12 weeks and 2&3 spacing 24 weeks, immunity conferred by dose 2&3 wanes to susceptibility after 26 weeks; (h) Dose 1&2 spacing 3 weeks and 2&3 spacing 8 weeks, immunity conferred by dose 2&3 wanes to susceptibility after 26 weeks.

We find consistent results in all scenarios that administration of booster dose reduces the ratio of cases and severe infections in the medium term up to 3 years (see Fig. 2 and Supplementary Fig. S2). For a variety of metrics of dose spacing, the outcomes do not fluctuate qualitatively under diverse hypotheses on dose immunity protection, administration rate, rollout initiation, and transmission traits of the pathogen. The allocation of a booster dose contributes positively to the mitigation and depletion of the pandemic. Attributable to the dynamics of transmission, an immediate mitigating or diminishing effect resulting from booster dose is not observed, especially for the initial weeks from the point of initiation. Rather, the effect manifests at later times in terms of lessening the peak size of the subsequent waves of the outbreak. Even if the booster dose is administered using a shortened 8-week dose spacing and implemented prior to the saturation, salient and immediate reduction of outbreak size does not occur. Speedier rollout of dose 3 (0.05 per week) observes a more salient effect, depleting more sharing of incidences and severe infections relative to slower rollout (0.02 per week). In the case where the rollout initiation of dose 1 is delayed for 2 weeks, the size of the outbreak will be enlarged approximately from 3.5% to 4.0%(Fig. 2c vs. 2e), which implies that the timing of primary vaccination can fundamentally affect the trajectory of transmission. Generally, the administration of booster vaccination is expected to induce a noticeably meaningful containment effect versus the primary two-dose strategy in terms of downgrading the saturation size of the pandemic at later times. Deployment of primary doses at an early stage is preferable, in contrast, early distribution of booster dose cannot facilitate immediate mitigation effect of outbreak saturation.

### Improving the coverage or administration capacity of primary vaccine series curtails cases and severe infections more versus booster vaccination strategy

We thereafter employ prior three-dose strategy (administration rate 0.02 per week) as the baseline and evaluate the performance of varying scenarios enhanced from primary vaccination strategy to estimate whether efficiency improvement of primary dose series can induce more desirable benefits when dose inadequacy or imbalance is of concern. Case 1 accelerates the rollout rate of dose 1 and doses 2 by 30% respectively; case 2 boosts gradually the administration rate of primary dose 1 over time; and case 3 doubles the administration rate of dose 1. In all these three settings, booster dose is not distributed.

We find that broadening the coverage or improving the capacity of primary vaccination series by accelerating rollout rate to vulnerable individuals who are previously unvaccinated reduces the ratio of total incidences and severe infections more versus the administration of booster vaccination to individuals who are vaccinated with primary doses (Fig.3a-3h and Supplementary Fig.S3) contingent on assumptions of dose spacing, the strength of waning immunity, initiation of administration and the traits of the pathogen. Asymptotically, doubling the administration rate of dose 1 yields the greatest downsize effect, followed by time-increment administration of dose 1 and then 30% growth of administration rate for dose 1&2 in all scenarios. The outcome does not change qualitatively across an extensive range of assumptions. It needs to be noted that a more salient and immediate mitigation effect in terms of peak downsize of the pandemic is observed during the initial weeks than later waves. Asymptotically, doubling the administration rate of dose 1 is expected to reduce the peak size of incidences from 4.0% to 1.9% versus booster vaccination, nearly 2.1% of improvement, then 1.2% decrement of size for time-increment rate enhancement, and 0.8% for the 30% growth of administration rates. The efficiency-improved primary vaccination strategy can fulfill meaningful and consistent containment effects across concurrent and later waves. The timing of primary dose 1 exerts a more salient depletion effect than the timing of booster dose 3 at the population level. Prioritizing primary doses instead of booster doses to more unvaccinated populations can induce more sizeable containment effects than the booster strategy. Consequently, vaccinating as many individuals with primary doses as possible needs to be taken with priority in settings when the insufficiency or imbalance of vaccines is of challenge. The finding supports the recommendation that prompt provision of primary doses to unprotected and vulnerable individuals outweighs the prioritized provision of booster vaccination to primary-dose vaccinated individuals. The containment effect resulting from vaccination capacity improvement in terms of rate increment is meaningful and sizeable even if the approach is enacted gradually. Hence, in settings where initially vaccine shortage is being challenged, it is of importance to give priority to the administration of the first dose or primary series to impart partial immunity at the population level rather than the prioritization of the booster dose. And if dose supply is promoted over time thereafter, it can be better off by increasing the availability of and accelerating the administration rate of primary doses to the population including individuals who are not previously vaccinated and individuals who have not completed the primary dose series.

**Fig 3.**
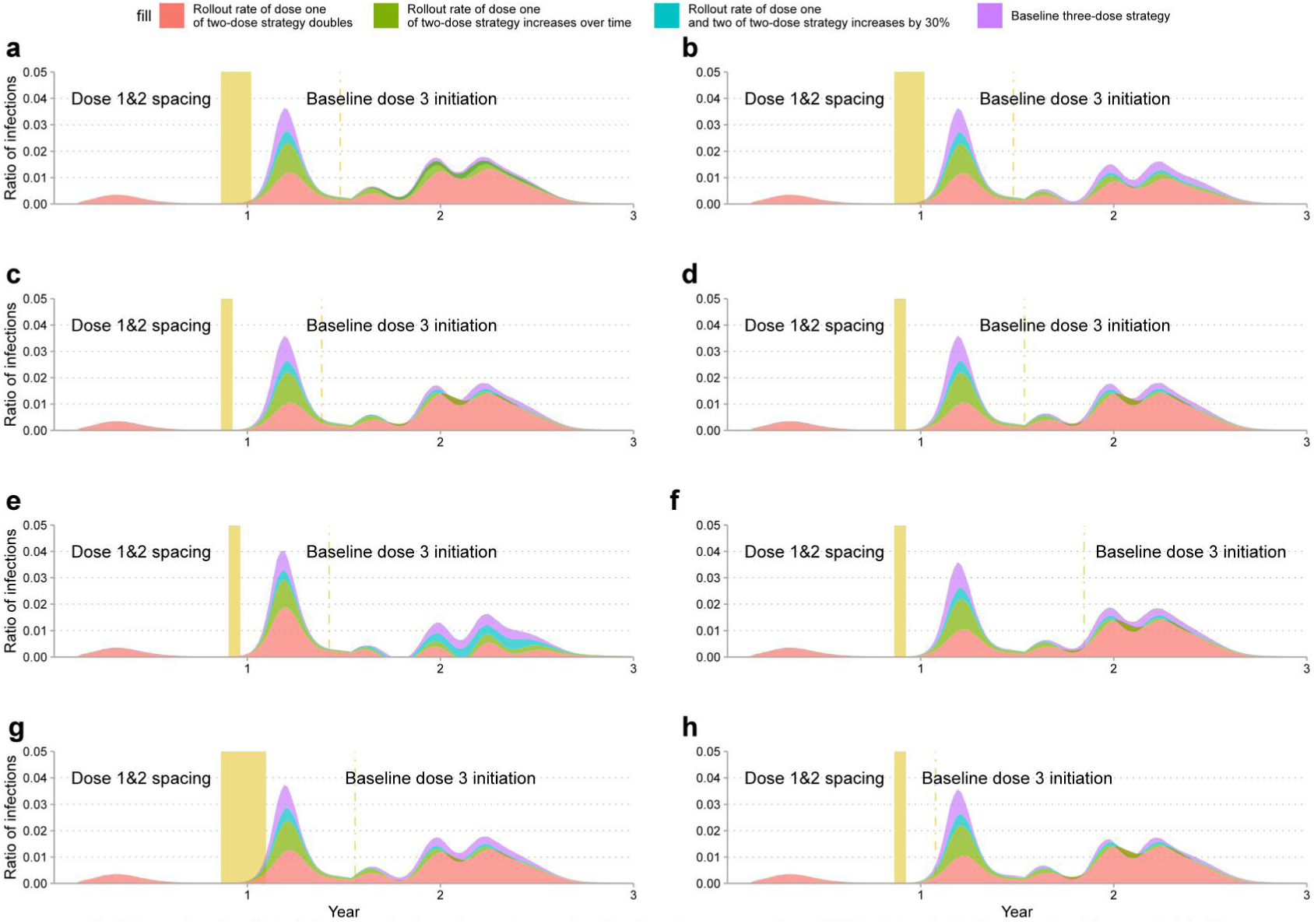
Comparison the effect of primary vaccination refinement versus baseline three-dose strategy. Dose 1 initiated at week 45 after the outbreak in scenario (a)∼(d) and (f)∼(h). Initiation of dose 1 in scenario (e) delayed to week 47.The immunity conferred by dose 1 wanes to susceptibility after 6.5 weeks in all scenarios. (a) Dose 1&2 spacing 8 weeks and 2&3 spacing 24 weeks, immunity conferred by dose 2&3 wanes to susceptibility after 26 weeks; (b) Dose 1&2 spacing 8 weeks and 2&3 spacing 24 weeks, immunity conferred by dose 2&3 wanes to susceptibility after 52 and 26 weeks respectively; (c) Dose 1&2 spacing 3 weeks and 2&3 spacing 24 weeks, immunity conferred by dose 2&3 wanes to susceptibility after 26 weeks; (d) Dose 1&2 spacing 3 weeks and 2&3 spacing 32 weeks, immunity conferred by dose 2&3 wanes to susceptibility after 26 weeks; (e) Dose 1&2 spacing 3 weeks and 2&3 spacing 24 weeks, immunity conferred by dose 2&3 wanes to susceptibility after 26 weeks; (f) Dose 1&2 spacing 3 weeks and 2&3 spacing 48 weeks, immunity conferred by dose 2&3 wanes to susceptibility after 26 weeks; (g) Dose 1&2 spacing 12 weeks and 2&3 spacing 24 weeks, immunity conferred by dose 2&3 wanes to susceptibility after 26 weeks; (h) Dose 1&2 spacing 3 weeks and 2&3 spacing 8 weeks, immunity conferred by dose 2&3 wanes to susceptibility after 26 weeks.

### Improving the administration rate of primary vaccination is feasible to render reproduction number below one that ultimately eradicates the pandemic in the absence of severe variants

The reproduction number of an infection is the expected number of cases directly generated by one index case in a population and mathematically denoted as *R* ^17^. If *R* > 1, the spread of pandemic can not be successfully contained; in contrast, if *R* < 1 the pandemic is to diminish ultimately (refer methods and supplementary material) ^17,27^. The size *R* reflects the transmission dynamics and severity of a pandemic and hinges on factors including the rate of vaccination, rate of immunity waning, transmission rate, rate of recovery, rate of infection, and susceptibility. Given the uncertainty of confounding factors regulating the transmission dynamics, we perform tests on vaccination rate and vaccine efficacy as well as principal parameters in shaping the epidemic, we assume dose 2 loses protection over time and uniformly wanes and the efficacy of dose 1 uniformly declines at three paces: (1) loses protection after 5 weeks; (2) loses protection after 13 weeks; and (3) loses protection after 26 weeks (Fig. 4). The infection risk after being vaccinated with dose 3 varies to account for potential heterogeneity. All other parameters used in the simulation are sketched in detail (See Supplementary Table S3). We assess the effect of vaccination rate on the size *R* contingent on the aforementioned parameters. The rate of booster dose is close to boundary to mimic the scenario that only small portion of individuals such as essential healthcare workers and immunity-compromised patients are vaccinated to obtain the necessary enhanced immunity. In the case where the likelihood of severe variants emergence and being infected is high, it can be of difficulty to render *R* below one even if healthcare service and administration rate is significantly promoted, rendering the containment of pandemic more long-lasting and costly (Fig. 4a). In settings where rollout rates are close to the boundary and healthcare service is not at equilibrium, it is of hardship to achieve *R* <1 to meet the criteria of depletion. Consistently, we find that the rollout rate is negatively correlated with the size of the reproduction number controlling for other factors. Upon constant administration of dose 2, increasing the rollout speed of dose 1 is expected to shape a reduced size of *R*. The outcome is consistent across a variety of vaccine protection profiles. Generally, the speedier rate of vaccination, the smaller magnitude of *R* and therefore the more expeditious containment of the pandemic. It is of importance to administer doses to the general population in a rate-prioritized way when conditions are met. Under the assumption of the commensurate magnitude of *R*, we observe a negative relationship between dose administration rates. Hence, to contain the exponential-like spreading resulting from initial administration inefficiency (e.g., a low administration rate), it is crucial to speed up the rollout of doses at a later time to ensure coverage of more individuals timely.

**Fig 4.**
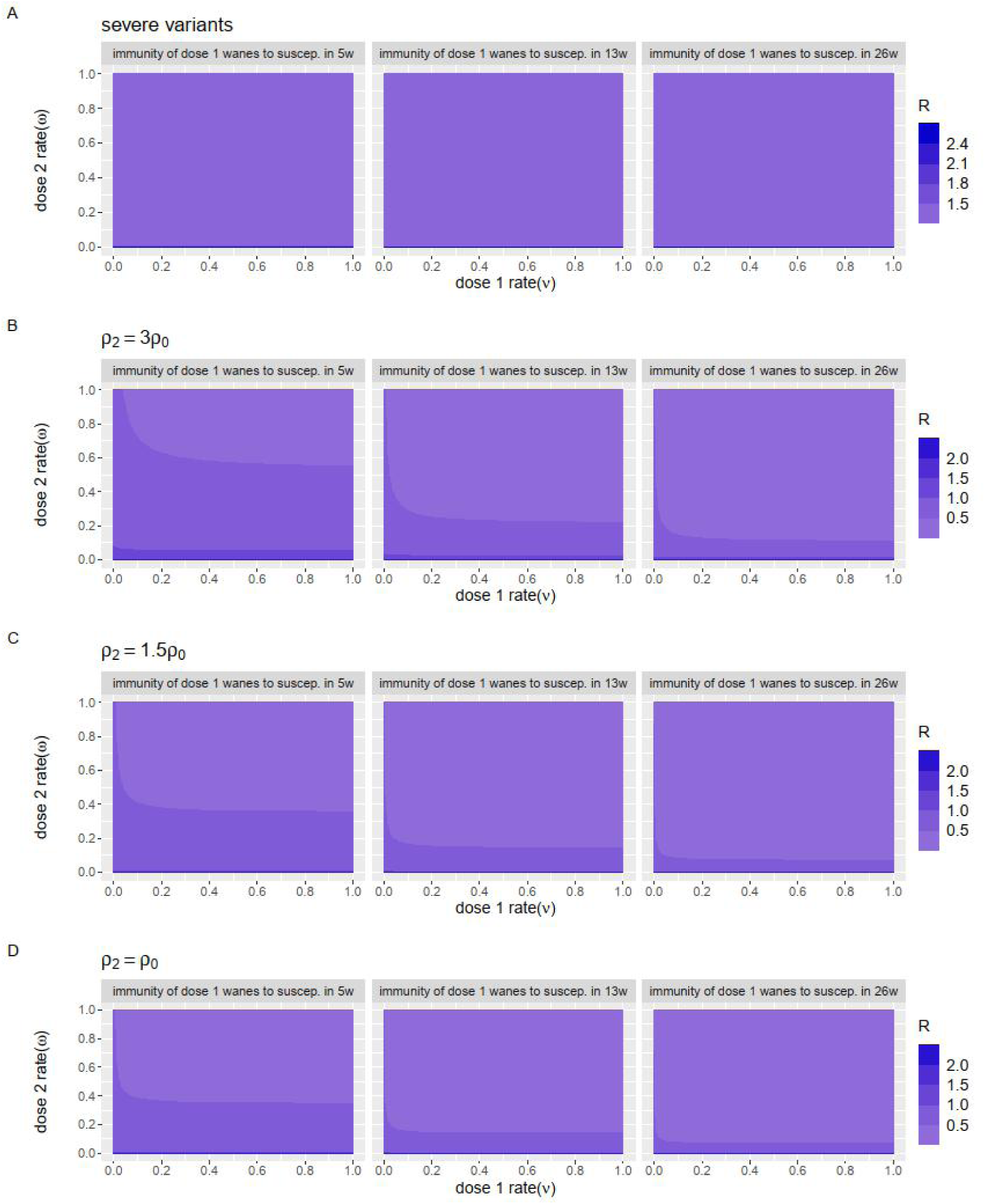
Impact of vaccination rate on the reproduction number of COVID-19 (A) Case where severe variant occurs;(B) Immunity of dose 2 & 3 wanes in 26 weeks; (C) Immunity of dose 2 & 3 wanes in 52 weeks; (B) Immunity of dose 2 & 3 wanes in 78 weeks;

Following the similar vein, we further evaluate the impact of the interaction of rollout rate on the epidemic size in terms of reproduction number. We assume dose 2 wanes evenly at three differentiated paces: (1) wanes immunity in 6 months; (2) wanes immunity in 12 months; and (3)wanes immunity in 18 months. Similar findings are identified across a variety of analyses (Fig. 5). All other parameters used in the simulation are illustrated in detail (See Supplementary Table S4). Inverse relationship is observed between the rates of administration. Under constant administration of dose 1, the growth of dose 2 administration rate ceteris paribus is associated with reduced size of reproduction number *R* in all cases. And efficiency-enhanced distribution of doses 1 is feasible to reduce the size of pandemic where the reproduction number *R* <1 under varying assumptions of dose immunity. By improving the administration capability of doses, it is capable of reducing and even diminishing the transmission essentially.

**Fig 5.**
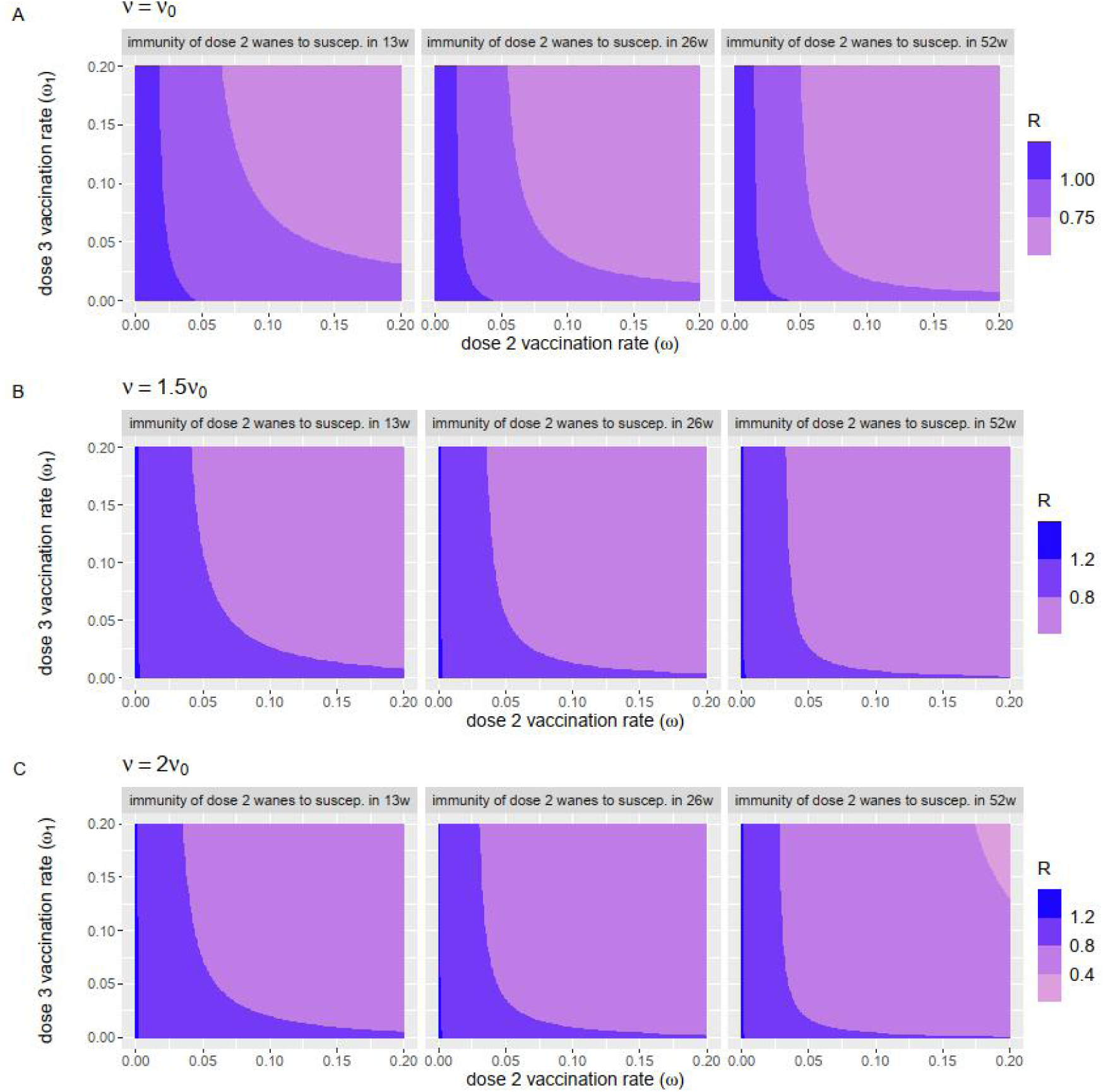
Impact of vaccination rate interaction on the dynamic reproduction number of COVID-19 pandemic. In all scenarios, Immunity of dose 2 & 3 wanes to susceptibility in 26 and 52 weeks respectively.

## Discussion

Our findings imply that vaccination strategy can considerably impact the trajectory of the COVID-19 epidemic. The contribution of our study is multi-fold. First, we show asymptotically by mathematical modeling and analysis that administrating booster dose to previously vaccinated individuals scales down the peak incidences of cases and severe infections in the absence of severe variants. The mitigation effect is salient at later waves rather than the immediate concurrent wave of the outbreak. Second, the administration of booster dose is suboptimal versus enhancing the rollout efficiency of primary vaccination in settings where dose inadequacy or imbalance is challenged and unvaccinated individuals remain to be of public concern. Increasing the availability of and administration rate of primary dose series for more vulnerable individuals can lower the total incidences and severe diseases more versus prioritizing the booster vaccination in both concurrent wave and subsequent outbreaks. The earlier rollout of primary doses outperforms delayed administration in terms of reduction of peak size controlling for confounding factors. The benefits increase with the rate at which vaccines are administered. Third, the findings are robust to diversified dose spacing and other factors, and the benefits of enhancing primary vaccination hold across a variety of assumptions. Fourth, although the transmission dynamics of a pandemic is potentially affected by complex factors, it is feasible to obtain reproductive number below one by accelerating the vaccination rate including primary doses. Hence, it could be better off by expanding the availability and administration rate of primary doses in settings where dose availability per individual is low. The benefits of primary vaccination outweigh the benefits of booster vaccination over an expansive range of analyses, the mitigation effect resulting from the efficiency-enhanced primary vaccination is considerable and meaningful regardless of whether the administration efficiency is corrected at a time-invariant or time-varying pace. Our finding supports the recommendation that priority be calibrated to the distribution of primary doses to the more vulnerable population not yet completing the primary sets as rapidly and early as possible. Nonetheless, the mass vaccination of booster dose may potentially represent an alternative to targeted vaccination strategies in the case where global vaccine supply capacity and imbalance are not of appreciable challenge.

Our findings are consistent with recommendations stating that immunogenicity of the primary vaccination is effective in the prevention of infection ^5^ and delaying the booster dose was beneficial to further reduce the rates of carriage ^24^. We examined the connection between the weekly vaccination speed, infectiousness, and immunity protection in determining the size of the outbreak and therefore potential optimal vaccination strategies. We found that differences in dose spacing are negligible when the weekly administration capacity is not close to boundary-binding and modestly high relative to the reproduction number (Fig. 4 and Fig. 5). Observational data has identified that one dose of COVID-19 vaccine was efficacious and safe for individuals to protect against symptomatic COVID-19 and was effective against severe disease ^30,31^. Even if sustained immunity is achieved after infection, observational evidence suggests that more than one-half of a general population needs to be vaccinated to attain herd immunity ^30^. Prioritizing the primary doses facilitates the progress of herd immunity and is preferable to booster vaccination consistently across varying scenarios accounting for differentiated administration rates, the transmissibility of the pathogen, and efficacy of the vaccines. Prioritization of booster dose was a suboptimal strategy in settings where dose insufficiency or imbalance is observed. Instead, prioritizing the availability of primary vaccination sets to more populations who are underserved was associated with a larger reduction in the peak size of total infections and severe outcomes. Under a strategy in which vaccines are retained for presently vaccinated individuals, this can potentially provide meaningful insights under the assumption of supply shortage.

Prior work suggests that timing of vaccination rollout and prioritizing first doses over others is expected to engender a sizable effect on curtailing mortality rate ^32^. The timing of vaccination relative to the epidemic establishment plays a crucial role in determining the effect on the outbreak. If vaccination delay emerges before an epidemic starts to unfold, the effectiveness of vaccination strategies is compromised. We extend the findings to booster dose coupled with the incidence of cases and severe infections. Although the provision of primary doses needs to be taken with priority versus booster vaccination, the outcomes are sensitive to the timing of administration. This highlights the necessity of timely and early delivery of primary vaccination to the vulnerable and needed individuals.

Some studies showed that the rate at which the COVID-19 vaccines advanced to the general population is critical ^22^. Others have identified the spillover effect that high vaccination rates were associated with lower infection rates at later times for the unvaccinated population ^33^. It has been found that approximately 60% of the supplied vaccines were administered in some countries. The compromised pace of vaccination affects the effect of containment ^18^. Hence, the importance and necessity of promoting administration efficiency could be manifold. To increase the rollout capacity, several issues including misinformation, vaccine hesitancy resulting from adverse effects, safety concerns, and compromised public confidence (e.g., frequent updates of vaccination recommendations) have to be addressed. These dilemmas have complicated the advance of vaccination campaigns globally ^18^. Although a safe and effective vaccine holds the greatest promise for controlling the pandemic, hesitancy to accept vaccines remains ^34^. Vaccine hesitancy, defined as the delay in the acceptance or denial of vaccination regardless of the positive outcomes of vaccination, complicates the containment of the pandemic. Some adverse effects can be general whereas others are individual-specific varying from vaccine to vaccine ^5^. Studies found that adverse events including injection site pain, fatigue, headache, and chills were reported in nearly one-third of vaccine recipients ^14^. To reduce vaccine hesitancy, identifying adverse events and rebuilding the public confidence toward vaccination is indispensable ^35^. Research so far documented that the acceptance rate of COVID-19 vaccine was up to 80% in some countries ^21,36^. Vaccine hesitancy remains a non-negligible barrier to meaningful containment of COVID-19. The challenge could be elaborate and context-specific, differentiating over time, communities, vaccines, which has been recognized as a hurdle in both poor and non-poor countries ^37^. Investigations targeting healthcare staff who were involved in the direct exposure risk of COVID-19 found that the likelihood of refusing vaccination was lower relative to their counterparts ^36^. Addressing the challenge of global vaccine hesitancy is an initial step in the efforts of vaccination programs ^38^. It is detrimental to the consolidation of immunization efforts and eradication of the pandemic ^39^. Persistent vaccine hesitancy can deteriorate the situation where unvaccinated population increases, which would potentially render the emergence of novel or severe variants as the likelihood of transmission and cross-transmission in these settings are higher ^19^. Recent variants such as Alpha, Delta, and Omicron compromised the immunity protection conferred by vaccines ^10^, and a more than 20-fold escape from vaccine-elicited neutralization by Omicron has been reported ^40,41^. Identification of Omicron variant in Africa and thereafter fast spreading to many other countries reflected the novel challenge and the latent risk ahead. Vaccines are expected to provoke spillover benefits to the public health and the healthcare system if they are accessible for the general public especially for vulnerable and under-served populations ^42^.

Not only has the COVID-19 pandemic impacted the routine primary immunization services worldwide, but also the booster vaccination. The debate on COVID-19 vaccine equity and priority remained ever since. The administration of a third dose is motivated by concerns of variants and immunity waning. Consistent findings on compromised vaccination services highlighted the global pervasiveness of disruption of essential health services and the urgency of improvement ^43^. In settings where dose supply is being challenged, prioritizing vulnerable individuals who are at appreciable risk and receipt of no vaccine would render greater benefits than otherwise. Partial gain might be obtained from booster vaccination, however, it will not outperform the benefits of prioritizing partial protection to the most vulnerable and unvaccinated currently. If vaccines were deployed where they need the most, they could advance the eradication of the pandemic by inhibiting the risk of more variants. WHO has called for a moratorium on boosting until the benefits of primary vaccination have been made available to more populations globally ^19^. Even with a limited protective effect, prioritizing primary vaccination can reduce total cases and severe outcomes ^44,45^. Although booster dose might not be the optimal option for the general population presently, it might be of necessity as the extension of primary series for specific target populations, immunocompromised individuals, and older individuals particularly, whose risk of infection is higher^46,47^. Vaccine booster dose policy decisions need to be based on evidence of benefits both for individuals and public health and obligations to secure global equity in vaccine access as a mechanism to minimize health impacts and transmission, and thereby reduce the risk of variants and prolongation of the pandemic^48,49^. The vast majority of current infections and COVID-19 cases are observed in unvaccinated individuals. New waves of variants were exacerbating the public health crisis worldwide ^50,40^. Although the option of further reducing the infections by enhancing immunity in vaccinated individuals is appealing, decisions for this purpose need to be evidence-based and consider the benefits and risks for both individuals and society ^41,51^. The decision to recommend is complex and requires beyond clinical and epidemiological observation, reflecting consideration of strategic priorities. In the current context, priority needs to be given to the protection of more vulnerable populations and the prevention of recurrence of cyclical outbreaks. Policymakers need to establish clear criteria for applying booster vaccines in the population. The decision needs to be based on immunological considerations, public confidence, vaccine hesitancy, side effects, vaccine availability and coupled with evaluations on specific disease control objectives ^50^.

Vaccine efficacy has been identified by observation data, but enhanced and continuous surveillance is still required as long-term observation of the dynamic is extraordinarily indispensable to inform efficacious countermeasures of the pandemic. More evaluations regarding the safety profile, immunogenicity, vaccine doses effect, and vaccination regimes are of necessity to minimize adverse events ^23^. Prompt deployment, equitable accessibility, and optimization of vaccination regimens to those who need the most, all of which are preliminary to the efforts of vaccination programs. Successful delivery of these efforts would contribute positively to the eradication of the pandemic and resume back to normal activities globally ^47^. Countries might forgo prioritizing the first doses approach due to concern about lacking supporting clinical data or other reasons ^32^. Lessons from other epidemics could provide insights and be integrated closely with communities and existing services that are familiar and readily accessible to local residents to improve the incentives of vaccination ^18^.

Globally, the overall supply of vaccines is growing but yet evenly distributed across countries. Lower-income countries suffer less accessibility, and vaccination is unpredictable and irregular ^2^. Considerations need to prioritize the coverage of primary vaccination series to ensure partial immunity in these settings. More data will be needed to understand the potential impact of booster vaccination on the duration and strength of protection against infection and transmission, particularly in the context of emerging variants. Public health and countermeasures continue to be an essential component of the COVID-19 prevention strategy, especially in light of the Omicron-like more infectious variants. In the global context of vaccine supply constraints and imbalance, the administration of booster doses potentially exacerbates vaccine access in countries with substantial vaccine coverage and diverting supply from settings that have not yet completed the primary vaccination. Particularly the currently available data show little evidence for widespread use of booster vaccination at population level ^51^. Literature verified that the primary-dose intervention strategy could decrease the size of pandemic ^17,49^.

COVID-19 vaccines will shape post-pandemic epidemiological trajectories and characteristics of immunity. It is therefore imperative to determine the strength and duration of protection and transmission-blocking immunity through thorough clinical evaluations to enforce sound public policies ^17^. Gaps remain in scientific understanding of the pathogen, and uncertainty regarding COVID-19 persists ^22^.

It is of importance to note that the results of our study are derived with the assumption of R = 2.3. Hence, in settings where the dynamics of epidemic unfolds following a disparate pathway and where the pathogen and vaccine present diversified traits assumed, the findings need to be calibrated and refined. To reasonably interpret the findings, it is crucial to reflect the limitations inherent in the analysis. Although we partially take into account the dynamics of the transmission by calibrating a time-varying case, most of the outcomes are based on deterministic models and time-invariant analysis, which might be disparate from the stochastic variability in practice. We conducted an extensive sensitivity analysis to show the degree of robustness by incorporating potential confounding parameters. The unvaccinated populations are accounted for in the model, but we did not explicitly rationalize the potential heterogeneity within these populations. Further stratified analysis may lead to refined findings. Although administration of booster dose at the population level is not of priority currently subject to resource limitations, it could be preferable to distribute locally booster dose at the individual level with priority to immunity-compromised patients and essential healthcare workers whose risk of being infected is considerably high. Analysis indicated that antibodies against COVID-19 conferred by vaccines were critical in disease prevention and recovery. Hence, long-term tracking the dynamics of immunity can provide important insight, prognosis, and control of COVID-19 ^46^. Further studies are required to understand whether heterogeneity of vaccination campaigns has rendered stratified containment effects. The supply of vaccines is increasing over time but global supply imbalance remains to be the challenge, the dynamics of prioritizing vaccination strategy is to update as well. Evidence is accumulating to inform global recommendations of strategy, which may be refined as more required data become available.

Our model-based evaluation highlights the merits of primary vaccination strategies in simultaneously optimizing containment objectives (e.g., the incidence of cases and severe infections). While there remains great uncertainty regarding the effectiveness of vaccines, especially differences in vaccine types, levels of service provision, and immunity waning dynamics, there is evidence to suggest that the way vaccines are administered is crucial. Finally, the simulation presented in this study can asymptotically provide insights for other countries to identify optimized vaccine strategies conditional on the availability of vaccines, stratified containment objectives, vaccine-specific traits, dynamics of transmission, administration capacity, the capacity of healthcare, public confidence, vaccine hesitancy and other potential factors in practical implementation. The effectiveness of a COVID-19 vaccine will be shaped by the efforts on delivering as quickly to the populations as possible ^53^. Coverage of vaccination to more populations to ensure partial immunity rather than otherwise in the current context would considerably impact the trajectory of the pandemic. The pathogen will not cease mutation before the timely administration of effective, affordable, and available vaccines to the public. When it is of difficulty to pinpoint the next outbreak, the prioritization the world is taking as of today would be of relevance ^52^.

## Methods

### Compartmental models

The model, schema, and assumptions employed in this study share the same principal immuno-epidemiological structure (Fig. 1 and Fig. S1) proposed by Saad-Roy et al. ^17^, extending the principal model framework by incorporating the booster dose compartment, pathogen, and transmission traits. The compartment *S*_***P***_ denotes full susceptible individuals who are not previously infected; *S*_***S***_ stands for partially susceptible individuals recovered from COVID-19 infection but exposed to risk again. *I*_***P***_, and *I*_***S***_ represent primary and secondary infections respectively from the corresponding susceptibility compartment; 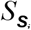 incorporates individuals vaccinated with dose *i* and thereafter immunity protection wanes over time; 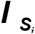 describes individuals being infected from compartment 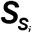; ***V***_*i*_ includes individuals vaccinated with dose *i*; *I*_*V*_ portrays individuals being infected from compartment ***V***_*i*_, and compartment *R* means recovery from the infection. *I*_*SR*_ denotes the incidence of severe infections of COVID-19. To allow for heterogeneity in immune protection, dose administration rate, and other potential confounding factors, we inherit epidemiological parameter estimates from available literature and summarize them in Supplementary Tables S1-S5. Given that estimates for the ratio of incidences and severe infections fluctuate, we allow these parameters to vary across a plausible range of scenarios.

## Supporting information

Supplemental File for Fig. S1-S3 and Table S1-S5

## Data Availability

No datasets are used in the analysis.

## Data availability

No datasets were utilized in this modeling study. Parameter calibrations for simulations were partially obtained from the literature and assumptions as described in Supplementary Table S1-S5.

## Code availability

All simulations and analyses were performed using R3.6.3, deSolve package1.28, and dplyr package 1.0.6. Code to run models presented in this paper is available upon reasonable request.

## Acknowledgments

This research was partly funded by the China Ministry of Education Industry and Education Cooperation Project [grant number 202002041005].

## Author contributions

I.N. conceptualized the study. I.N. designed models and implemented simulations, performed analyses, interpreted results and wrote the manuscript.

## References

1. World Health Organization. WHO Coronavirus (COVID-19) Dashboard. https://covid19.who.int (2022).

2. Wouters, O. J. et al. Challenges in ensuring global access to COVID-19 vaccines: production, affordability, allocation, and deployment. Lancet 397, 1023–34(2021).

3. World Health Organization. WHO COVID-19 vaccines https://www.who.int/emergencies/diseases/novel-coronavirus-2019/covid-19-vaccines (2022).

4. Chung. J. Y. et al. COVID-19 vaccines: The status and perspectives in delivery points of view. Adv. Drug. Deliv. Rev. 170, 1–25 (2021).

5. Meo, S. A. et al. COVID-19 vaccines: comparison of biological, pharmacological characteristics and adverse effects of Pfizer/BioNTech and Moderna Vaccines. Eur. Rev. Med. Pharmacol. Sci. 25, 1663–1669 (2021).

6. World Health Organization. WHO COVID-19 advice for the public. https://www.who.int/emergencies/diseases/novel-coronavirus-2019/covid-19-vaccines/advice (2022).

7. Mathieu, E. et al. A global database of COVID-19 vaccinations. Nat. Hum. Behav. 5, 947–953 (2021).

8. Our World in Data. Statistics and Research Coronavirus (COVID-19) Vaccinations.https://ourworldindata.org/covid-vaccinations (Accessed on January 5, 2022).

9. Harrison, E. A. et al. Vaccine confidence in the time of COVID-19. Eur. J. Epidemiol. 35, 325–330 (2020).

10. Jecker, N. S. et al. Three for me and none for you? An ethical argument for delaying COVID-19 boosters. J. Med. Ethics., 10.1136/medethics-2021-107824 (2021).

11. World Health Organization. Interim statement on booster doses for COVID-19 vaccination. https://www.who.int/news/item/22-12-2021-interim-statement-on-booster-doses-for-covid-19-vaccination---update-22-december-2021 (2022).

12. Polack, F. P. et al. Safety and Efficacy of the BNT162b2 mRNA Covid-19 Vaccine. N. Engl. J. Med. 383, 2603–15 (2020).

13. Vasileiou, E. et al. Interim findings from first-dose mass COVID-19 vaccination roll-out and COVID-19 hospital admissions in Scotland: a national prospective cohort study. Lancet 397, 1646–57(2021).

14. Lamb, Y.N. et al. BNT162b2 mRNA COVID-19 Vaccine: First Approval. Drugs 81, 495–501 (2021).

15. Sadoff, J. Safety and Efficacy of Single-Dose Ad26.COV2.S Vaccine against Covid-19. N. Engl. J. Med. 384, 2187–201 (2021).

16. Mehrotra, D. V. et al. Clinical Endpoints for Evaluating Efficacy in COVID-19 Vaccine Trial. Ann. Intern. Med. 10.7326/M20-6169 (2021).

17. Saad-Roy, C.M. et al. Epidemiological and evolutionary considerations of SARS-CoV-2 vaccine dosing regimes. Science 372, 363–370 (2021).

18. Mallapaty, S. Researchers fear growing COVID vaccine hesitancy in developing nations. Nature, 10.1038/d41586-021-03830-7(2021).

19. Krause, P. R. et al. Considerations in boosting COVID-19 vaccine immune responses. Lancet 398, 1377–80 (2021).

20. Voysey, M. et al. Safety and efficacy of the ChAdOx1 nCoV-19 vaccine (AZD1222) against SARS-CoV-2: an interim analysis of four randomised controlled trials in Brazil, South Africa, and the UK. Lancet 397, 99–111 (2021).

21. Troiano, G. Vaccine hesitancy in the era of COVID-19. Public Health 194, 245–251 (2021).

22. Verbeke, R. et al. The dawn of mRNA vaccines: The COVID-19 case. J. Control Release. 333, 511–520 (2021).

23. Yang, S.L. et al. Safety and immunogenicity of a recombinant tandem-repeat dimeric RBD-based protein subunit vaccine (ZF2001) against COVID-19 in adults: two randomised, double-blind, placebo-controlled, phase 1 and 2 trials. Lancet Infect. Dis. 21, 1107–19 (2021).

24. Harmonia, N. A. et al. Modelling the effects of booster dose vaccination schedules and recommendations for public health immunization programs: the case of Haemophilus influenzae serotype b. BMC Public Health 17, 705 (2017).

25. Experton, B. et al. A Predictive Model for Severe COVID-19 in the Medicare Population: A Tool for Prioritizing Primary and Booster COVID-19 Vaccination. Biology 10, 1185 (2021).

26. Katikireddi, S.V. et al. Two-dose ChAdOx1 nCoV-19 vaccine protection against COVID-19 hospital admissions and deaths over time: a retrospective, population-based cohort study in Scotland and Brazil. Lancet, 10.1016/S0140-6736(21)02754-9 (2021).

27. Bubar, K. M. et al. Model-informed COVID-19 vaccine prioritization strategies by age and serostatus. Science 371, 916–921 (2021).

28. Polack, FP. et al. Safety and efficacy of the BNT162b2 mRNA Covid-19 vaccine. N. Engl. J. Med. 383: 2603–2615 (2020).

29. Barda, N. et al. Effectiveness of a third dose of the BNT162b2 mRNA COVID-19 vaccine for preventing severe outcomes in Israel: an observational study. Lancet 2021, 398, 10316, 2093-2100 (2021).

30. Koirala, A. et al. Vaccines for COVID-19: The current state of play. Paediatr. Respir. Rev. 35, 43–49 (2020).

31. Halperin, S. A. Final efficacy analysis, interim safety analysis, and immunogenicity of a single dose of recombinant novel coronavirus vaccine (adenovirus type 5 vector) in adults 18 years and older: an international, multicentre, randomised, double-blinded, placebo-controlled phase 3 trial. Lancet, 10.1016/S0140-6736(21)02753-7 (2021).

32. Wang, X. et al. Effects of COVID-19 Vaccination Timing and Risk Prioritization on Mortality Rates, United States. Emerg. Infect. Dis. 27(7), 1976–1979 (2021).

33. Milman, O. et al. Community-level evidence for SARS-CoV-2 vaccine protection of unvaccinated individuals. Nat. Med. 27, 1367–1369 (2021).

34. Kaplan R. M. et al. Influence of a COVID-19 vaccine’s effectiveness and safety profile on vaccination acceptance. PNAS 118, 1–5 (2021).

35. Haynes, B. F. et al. Prospects for a safe COVID-19 vaccine. Sci. Transl. Med. 12, eabe0948 (2020).

36. Dror, A.A. et al. Vaccine hesitancy: the next challenge in the fight against COVID-19. Eur. J. Epidemiol. 35, 775–779 (2020).

37. Kate, S. et al. COVID-19 vaccines: rapid development, implications, challenges, and future prospects. Hum. Cell 34, 711–733 (2021).

38. Sallam, M. COVID-19 Vaccine Hesitancy Worldwide: A Concise Systematic Review of Vaccine Acceptance Rates. Vaccines 9, 160 (2021).

39. Wu, J. et al. COVID-19 Vaccine Hesitancy Among Chinese Population: A Large-Scale National Study. Front. Immunol. 12, 781161(2021).

40. Cele, S. et al. Omicron extensively but incompletely escapes Pfizer BNT162b2 neutralization. Nature, 10.1038/d41586-021-03824-5 (2021).

41. Planas, D. et al. Considerable escape of SARS-CoV-2 Omicron to antibody neutralization. Nature, 10.1038/d41586-021-03827-2 (2021).

42. Greter, H. et al. Heterologous vaccine regimen: Stakeholder acceptance and implementation considerations. Vaccine 39, 580–587 (2021).

43. Shet, A. et al. Impact of the SARS-CoV-2 pandemic on routine immunisation services: evidence of disruption and recovery from 170 countries and territories. Lancet Glob. Health. 2021, 10.1016/S2214-109X(21)00512-X.

44. Corey, L. et al. A strategic approach to COVID-19 vaccine R&D. Science 368, 948–950 (2020).

45. Worby, C. J. et al. Face mask use in the general population and optimal resource allocation during the COVID-19 pandemic. Nat. Commun. 11, 4049 (2020).

46. Li, K. et al. Dynamic changes in anti-SARS-CoV-2 antibodies during SARS-CoV-2 infection and recovery from COVID-19. Nat. Commun. 11, 6044 (2020).

47. Kim, J. H. et al. Looking beyond COVID-19 vaccine phase 3 trials. Nat. Med. 27, 205–211 (2021).

48. Arbel, R. et al. BNT162b2 Vaccine Booster and Mortality Due to Covid-19. N. Engl. J. Med. 385, 2413–20(2021).

49. Misra, O.P. et al. Modelling the effect of booster vaccination on the transmission dynamics of diseases that spread by droplet infection. Nonlinear Anal. Hybrid Syst. 3, 657–665 (2009).

50. Munro, A. P. S. et al. Safety and immunogenicity of seven COVID-19 vaccines as a third dose (booster) following two doses of ChAdOx1 nCov-19 or BNT162b2 in the UK (COV-BOOST): a blinded, multicentre, randomised, controlled, phase 2 trial. Lancet 398, 2258–76 (2021).

51. Krause, P. R. et al. Considerations in boosting COVID-19 vaccine immune responses. Lancet 398, 10308, 1377-1380 (2021).

52. Alagoz, O. et al. The impact of vaccination to control COVID-19 burden in the United States: A simulation modeling approach. PLoS ONE 16, e0254456 (2021).

53. Paltiel, A. D. et al. Clinical Outcomes Of A COVID-19 Vaccine: Implementation Over Efficacy. Health Aff. 40, 42–52 (2021).

