## Supplemental File for Fig. S1-S3 and Table S1-S5 for "Evaluation of the effects of vaccination regimes on the transmission dynamics of COVID-19 pandemic"

Ichiro Nakamoto

Supplementary Methods
Supplementary Figures S1-S3
Supplementary Table S1-S5
References

### Supplementary Methods

Suppose a susceptible-vaccinated-infected-recovered-susceptible[SVIRS] model and a population of size N, in which each individual can be vaccinated or unvaccinated (Fig. S1). Assume the birth rate of the population is equivalent to the death rate $\mu$ , individuals recover from infection at the rate $\gamma$ and the thereafter immunity wanes at rate $\delta$ after recovery. Full susceptible individuals get primary doses 1&2 vaccinated at rate $\nu$ and $\omega$ , and get the booster dose 3 vaccinated at rate $\omega_1$ respectively. The immunity protection of doses 1&2&3 declines at rate $\rho_1$ , $\rho_2$ and $\rho_3$ respectively. Vaccinated individuals whose immunity wanes are infected at rate $\varepsilon_1$, $\varepsilon_2$ and $\varepsilon_3$. $S_{vax}$ , $S_{vax_2}$ $S_{vax_3}$ and denote the administration initiation timing of doses 1, 2, and 3. $\varepsilon_{V_i}$ is the infection rate after vaccination of dose $i$ . Partially susceptible individuals can be vaccinated again and obtain the immunity protection equivalent to doses 1&2&3 at rate $c\nu$ , $d\nu$ and $(1-c-d)\nu$ . $\lambda_i$ is the likelihood of being severe disease due to the infection of COVID-19.

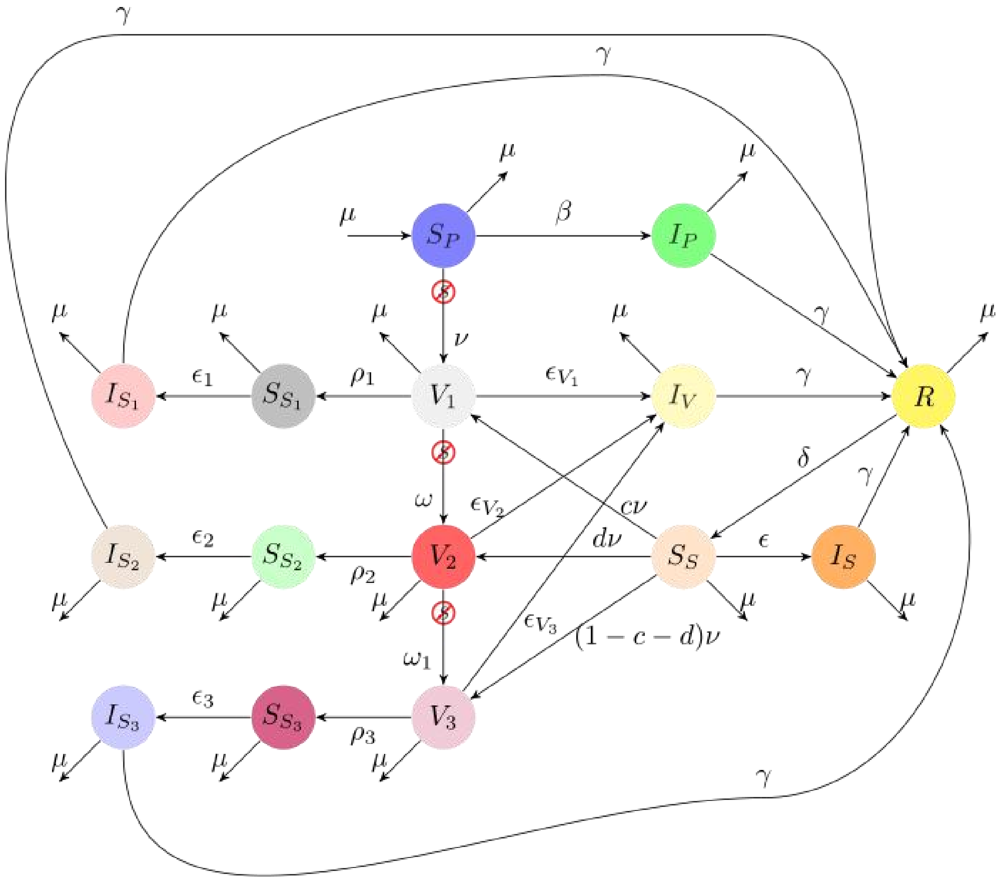


Figure S1: COVID-19 transmission dynamics

- Dose 1 vaccination
- Dose-1 immunity wanes
- Infection after dose-1 immunity wanes
- Dose 2 vaccination
- Dose-2 immunity wanes
- Primary infection
- Infection after vaccination
- Recovered
- Partial susceptibility
- Secondary infection
- Dose 3 vaccination
- Dose-3 immunity wanes
- Infection after dose-3 immunity wanes
- Full susceptibility
- Infection after dose-2 immunity wanes

The model governing the epidemic transmission dynamics can be expressed as follows:

$$\frac{dS_P}{dt} = \mu - \beta S_P\left[I_P + \alpha I_S + \alpha_V I_V + \alpha_1 I_{S_1} + \alpha_2 I_{S_2} + \alpha_3 I_{S_3}\right] - \left(s_{vax}\nu + \mu\right)S_P \qquad (1)$$

$$\frac{dI_P}{dt} = \beta S_P\left[I_P + \alpha I_S + \alpha_V I_V + \alpha_1 I_{S_1} + \alpha_2 I_{S_2} + \alpha_3 I_{S_3}\right] - \left(\gamma + \mu\right)I_P \qquad (2)$$

$$\frac{dR}{dt} = \gamma\left[I_P + I_S + I_V + I_{S_1} + I_{S_2} + I_{S_3}\right] - \left(\delta + \mu\right)R \qquad (3)$$

$$\frac{dS_S}{dt} = \delta R - \varepsilon\beta S_S\left[I_P + \alpha I_S + \alpha_V I_V + \alpha_1 I_{S_1} + \alpha_2 I_{S_2} + \alpha_3 I_{S_3}\right] - \left(s_{vax}\nu + \mu\right)S_S \qquad (4)$$

$$\frac{dI_S}{dt} = \varepsilon\beta S_S\left[I_P + \alpha I_S + \alpha_V I_V + \alpha_1 I_{S_1} + \alpha_2 I_{S_2} + \alpha_3 I_{S_3}\right] - \left(\gamma + \mu\right)I_S \qquad (5)$$

$$\frac{dV_1}{dt} = s_{vax}\nu S_P + c s_{vax}\nu S_S - \varepsilon_{V_1}\beta V_1\left[I_P + \alpha I_S + \alpha_V I_V + \alpha_1 I_{S_1} + \alpha_2 I_{S_2} + \alpha_3 I_{S_3}\right] -$$
$$\left(\omega s_{vax1} + \rho_1 + \mu\right)V_1 \qquad (6)$$

$$\frac{dV_2}{dt} = d s_{vax}\nu S_S + \omega s_{vax_2}V_1 - \varepsilon_{V_2}\beta V_2\left[I_P + \alpha I_S + \alpha_V I_V + \alpha_1 I_{S_1} + \alpha_2 I_{S_2} + \alpha_3 I_{S_3}\right] -$$
$$\left(\omega_1 s_{vax_2} + \rho_2 + \mu\right)V_2 \qquad (7)$$

$$\frac{dV_3}{dt} = (1 - c - d)s_{vax}\nu S_S + \omega_1 s_{vax3}V_2 - \varepsilon_{V_3}\beta V_3\left[I_P + \alpha I_S + \alpha_V I_V + \alpha_1 I_{S_1} + \alpha_2 I_{S_2} + \alpha_3 I_{S_3}\right] -$$
$$\left(\rho_3 + \mu\right)V_3 \qquad (8)$$

$$\frac{dI_V}{dt} = \beta(\varepsilon_{V_1}V_1 + \varepsilon_{V_2}V_2 + \varepsilon_{V_3}V_3)\left[I_P + \alpha I_S + \alpha_V I_V + \alpha_1 I_{S_1} + \alpha_2 I_{S_2} + \alpha_3 I_{S_3}\right] - \left(\gamma + \mu\right)I_V \qquad (9)$$

$$\frac{dS_{S_1}}{dt} = \rho_1 V_1 - \varepsilon_1\beta S_{S_1}\left[I_P + \alpha I_S + \alpha_V I_V + \alpha_1 I_{S_1} + \alpha_2 I_{S_2} + \alpha_3 I_{S_3}\right] - \mu S_{S_1} \qquad (10)$$

$$\frac{dS_{S_2}}{dt} = \rho_2 V_2 - \varepsilon_2\beta S_{S_2}\left[I_P + \alpha I_S + \alpha_V I_V + \alpha_1 I_{S_1} + \alpha_2 I_{S_2} + \alpha_3 I_{S_3}\right] - \mu S_{S_2} \qquad (11)$$

$$\frac{dS_{S_3}}{dt} = \rho_3 V_3 - \varepsilon_3\beta S_{S_3}\left[I_P + \alpha I_S + \alpha_V I_V + \alpha_1 I_{S_1} + \alpha_2 I_{S_2} + \alpha_3 I_{S_3}\right] - \mu S_{S_3} \qquad (12)$$

$$\frac{dI_{S_1}}{dt} = \varepsilon_1\beta S_{S_1}\left[I_P + \alpha I_S + \alpha_V I_V + \alpha_1 I_{S_1} + \alpha_2 I_{S_2} + \alpha_3 I_{S_3}\right] - \left(\gamma + \mu\right)I_{S_1} \qquad (13)$$

$$\frac{dI_{S_2}}{dt} = \varepsilon_2 \beta S_{S_2}\left[I_P + \alpha I_S + \alpha_V I_V + \alpha_1 I_{S_1} + \alpha_2 I_{S_2} + \alpha_3 I_{S_3}\right] - (\gamma + \mu) I_{S_2} \qquad (14)$$

$$\frac{dI_{S_3}}{dt} = \varepsilon_3 \beta S_{S_3}\left[I_P + \alpha I_S + \alpha_V I_V + \alpha_1 I_{S_1} + \alpha_2 I_{S_2} + \alpha_3 I_{S_3}\right] - (\gamma + \mu) I_{S_3} \qquad (15)$$

$$I_{SR} = \lambda_P I_P + \lambda_S I_S + \lambda_V I_V + \lambda_1 I_{S_1} + \lambda_2 I_{S_2} + \lambda_3 I_{S_3} \qquad (16)$$

$$S_{vax} = \begin{cases} 0, & t < t_{vax} \\ 1, & t \geq t_{vax} \end{cases} \qquad (17)$$

$$S_{vax1} = \begin{cases} 0, & t < t_{vax1} \\ 1, & t \geq t_{vax1} \end{cases} \qquad (18)$$

$$S_{vax2} = \begin{cases} 0, & t < t_{vax2} \\ 1, & t \geq t_{vax2} \end{cases} \qquad (19)$$

In a disease-free equilibrium, no incidence of infections occurs and thus the system can be rephrased to below:

$$\frac{dS_P}{dt} = \mu - (\nu + \mu) S_P = 0 \qquad (20)$$

$$\frac{dV_1}{dt} = \nu S_P - (\omega + \rho_1 + \mu) V_1 = 0 \qquad (21)$$

$$\frac{dV_2}{dt} = \omega \mathbf{V}_1 - (\omega_1 + \rho_2 + \mu) V_2 = 0 \qquad (22)$$

$$\frac{dV_3}{dt} = \omega_1 \mathbf{V}_2 - (\rho_3 + \mu) V_3 = 0 \qquad (23)$$

$$\frac{dS_{S_1}}{dt} = \rho_1 \mathbf{V}_1 - \mu S_{S_1} = 0 \qquad (24)$$

$$\frac{dS_{S_2}}{dt} = \rho_2 V_2 - \mu S_{S_2} = 0 \qquad (25)$$

$$\frac{dS_{S_3}}{dt} = \rho_3 V_3 - \mu S_{S_3} = 0 \qquad (26)$$

The solution for the disease-free equilibrium is derived as follows:

$$S_P^* = \frac{\mu}{\nu + \mu} \qquad (27)$$

$$V_1^* = \frac{\nu}{\omega + \rho_1 + \mu} \cdot \frac{\mu}{\mu + \nu} \qquad (28)$$

$$V_2^* = \frac{\omega}{\omega_1 + \rho_2 + \mu} \cdot \frac{\nu}{\omega + \rho_1 + \mu} \cdot \frac{\mu}{\mu + \nu} \qquad (29)$$

$$V_3^* = \frac{\omega_1}{(\rho_3 + \mu)} \cdot \frac{\omega}{(\omega_1 + \rho_2 + \mu)} \cdot \frac{\nu}{\omega + \rho_1 + \mu} \cdot \frac{\mu}{\mu + \nu} \qquad (30)$$

$$S_{S_1}^* = \frac{\rho_1}{\mu} V_1^* = \frac{\rho_1}{\omega + \rho_1 + \mu} \cdot \frac{\nu}{\mu + \nu} \qquad (31)$$

$$S_{S_2}^* = \frac{\rho_2}{\omega_1 + \rho_2 + \mu} \cdot \frac{\omega}{\omega + \rho_1 + \mu} \cdot \frac{\nu}{\mu + \nu} \qquad (32)$$

$$S_{S_3}^* = \frac{\rho_3}{(\rho_3 + \mu)} \cdot \frac{\omega_1}{(\omega_1 + \rho_2 + \mu)} \cdot \frac{\omega}{\omega + \rho_1 + \mu} \cdot \frac{\nu}{\mu + \nu} \qquad (33)$$

Accordingly, the reproduction number for the transmission dynamics system is governed by the formula (34):

$$\Re = \frac{\beta}{\gamma+\mu}\left[S_P + \varepsilon \mathbf{S}_S + \varepsilon_{V_1}V_1 + \varepsilon_{V_2}V_2 + \varepsilon_{V_3}V_3 + \varepsilon_1 S_{S_1} + \varepsilon_2 S_{S_2} + \varepsilon_3 S_{S_3}\right]$$

$$= \frac{\beta}{\gamma+\mu}\left\{ \begin{array}{l} \frac{\mu}{\nu+\mu}\left[ \begin{array}{l} 1 + \frac{\nu\varepsilon_{V_1}}{\omega+\rho_1+\mu} + \frac{\nu\varepsilon_{V_2}}{\omega+\rho_1+\mu}\frac{\omega}{\omega_1+\rho_2+\mu} + \\ \frac{\nu\varepsilon_{V_3}}{\omega+\rho_1+\mu}\frac{\omega}{\omega_1+\rho_2+\mu}\frac{\omega_1}{\rho_3+\mu} \end{array} \right] + \\ \frac{\nu}{\nu+\mu}\left[ \begin{array}{l} \frac{\rho_1\varepsilon_1}{\omega+\rho_1+\mu} + \frac{\rho_2\varepsilon_2}{\omega+\rho_1+\mu}\frac{\omega}{\omega_1+\rho_2+\mu} + \\ \frac{\rho_3\varepsilon_3}{\omega+\rho_1+\mu}\frac{\omega}{\omega_1+\rho_2+\mu}\frac{\omega_1}{\rho_3+\mu} \end{array} \right] \end{array} \right\} \quad (34)$$

The partial first-order differential concerning the administration rate of dose 1 is :

$$\frac{\partial \Re}{\partial \nu} = \frac{\beta}{\gamma+\mu}\left\{ \begin{array}{l} \frac{-\mu}{(\nu+\mu)^2}\left[ \begin{array}{l} 1 + \frac{\nu\varepsilon_{V_1}}{\omega+\rho_1+\mu} + \frac{\nu\varepsilon_{V_2}}{\omega+\rho_1+\mu}\frac{\omega}{\omega_1+\rho_2+\mu} + \\ \frac{\nu\varepsilon_{V_3}}{\omega+\rho_1+\mu}\frac{\omega}{\omega_1+\rho_2+\mu}\frac{\omega_1}{\rho_3+\mu} \end{array} \right] + \\ \frac{\mu}{(\nu+\mu)^2}\left[ \begin{array}{l} \frac{\rho_1\varepsilon_1}{\omega+\rho_1+\mu} + \frac{\rho_2\varepsilon_2}{\omega+\rho_1+\mu}\frac{\omega}{\omega_1+\rho_2+\mu} + \\ \frac{\rho_3\varepsilon_3}{\omega+\rho_1+\mu}\frac{\omega}{\omega_1+\rho_2+\mu}\frac{\omega_1}{\rho_3+\mu} \end{array} \right] \end{array} \right\} \quad (35)$$

The sign of $\frac{\partial R}{\partial \nu}$ is indeterministic and hinges on multiple factors including dose administration rate, dose immunity waning rate, infection rate after vaccination, birth rate, and death rate. For reproduction number to be a monotonic decreasing function of the vaccination rate of dose 1

(i.e., $\frac{\partial R}{\partial \nu} < 0$), the condition of below needs to be met:

$$(\omega_1+\rho_2+\mu)(\rho_3+\mu)(\rho_1\varepsilon_1-\nu\varepsilon_{V_1})+(\rho_3+\mu)\omega(\rho_2\varepsilon_2-\nu\varepsilon_{V_2})+\omega\omega_1(\rho_3\varepsilon_3-\nu\varepsilon_{V_3})<$$
$$(\omega+\rho_1+\mu)(\omega_1+\rho_2+\mu)(\rho_3+\mu) \qquad (36)$$

After reorganizing and simplification, we obtain the condition as stated in formula (37):

$$\frac{(\rho_1\varepsilon_1-\nu\varepsilon_{V_1})}{(\omega+\rho_1+\mu)}+\frac{\omega(\rho_2\varepsilon_2-\nu\varepsilon_{V_2})}{(\omega+\rho_1+\mu)(\omega_1+\rho_2+\mu)}+\frac{\omega\omega_1(\rho_3\varepsilon_3-\nu\varepsilon_{V_3})}{(\omega+\rho_1+\mu)(\omega_1+\rho_2+\mu)(\rho_3+\mu)}<1 \qquad (37)$$

Following the similar vein as dose 1, we can derive the relative relation over the administration rate of dose 2:

$$\frac{\partial\Re}{\partial\omega}=\frac{\beta}{\gamma+\mu}\left\{\begin{array}{l}\dfrac{\mu}{\nu+\mu}\left[\begin{array}{l}\dfrac{-\nu\varepsilon_{V_1}}{(\omega+\rho_1+\mu)^2}+\dfrac{\nu\varepsilon_{V_2}}{(\omega+\rho_1+\mu)^2}\dfrac{\rho_1+\mu}{\omega_1+\rho_2+\mu}+\\ \dfrac{\nu\varepsilon_{V_3}}{(\omega+\rho_1+\mu)^2}\dfrac{\rho_1+\mu}{\omega_1+\rho_2+\mu}\dfrac{\omega_1}{\rho_3+\mu}\end{array}\right]+\\ \dfrac{\nu}{\nu+\mu}\left[\begin{array}{l}\dfrac{-\rho_1\varepsilon_1}{(\omega+\rho_1+\mu)^2}+\dfrac{\rho_2\varepsilon_2}{(\omega+\rho_1+\mu)^2}\dfrac{\rho_1+\mu}{\omega_1+\rho_2+\mu}+\\ \dfrac{\rho_3\varepsilon_3}{(\omega+\rho_1+\mu)^2}\dfrac{\rho_1+\mu}{\omega_1+\rho_2+\mu}\dfrac{\omega_1}{\rho_3+\mu}\end{array}\right]\end{array}\right\} \qquad (38)$$

For R to be a monotonic decreasing function of dose 2 administration rate, it needs to satisfy the condition $\frac{\partial R}{\partial \omega} < 0$:

$$(\mu\varepsilon_{V_2} + \rho_2\varepsilon_2)(\rho_1 + \mu)(\rho_3 + \mu) + (\mu\varepsilon_{V_3} + \rho_3\varepsilon_3)(\rho_1 + \mu)\omega_1 <$$
$$(\mu\varepsilon_{V_1} + \rho_1\varepsilon_1)(\omega_1 + \rho_2 + \mu)(\rho_3 + \mu) \qquad (39)$$

And after simplification, we obtain the condition of (40):

$$\frac{\mu\varepsilon_{V_2} + \rho_2\varepsilon_2}{(\rho_2 + \mu)(\omega_1 + \rho_2 + \mu)} + \frac{(\mu\varepsilon_{V_3} + \rho_3\varepsilon_3)\omega_1}{(\rho_3 + \mu)(\rho_2 + \mu)(\omega_1 + \rho_2 + \mu)} < \frac{\mu\varepsilon_{V_1} + \rho_1\varepsilon_1}{(\rho_1 + \mu)(\rho_2 + \mu)} \qquad (40)$$

Generalize the analysis to dose 3:

$$\frac{\partial \Re}{\partial \omega_1} = \frac{\beta}{\gamma + \mu}\left\{ \begin{array}{l} \frac{\mu}{\nu + \mu}\left[ \begin{array}{l} \frac{\nu\varepsilon_{V_2}}{\omega + \rho_1 + \mu}\frac{-\omega}{(\omega_1 + \rho_2 + \mu)^2} + \\ \frac{\nu\varepsilon_{V_3}}{\omega + \rho_1 + \mu}\frac{\omega}{(\omega_1 + \rho_2 + \mu)^2}\frac{\rho_2 + \mu}{\rho_3 + \mu} \end{array} \right] + \\ \frac{\nu}{\nu + \mu}\left[ \begin{array}{l} \frac{\rho_2\varepsilon_2}{\omega + \rho_1 + \mu}\frac{-\omega}{(\omega_1 + \rho_2 + \mu)^2} + \\ \frac{\rho_3\varepsilon_3}{\omega + \rho_1 + \mu}\frac{\omega}{(\omega_1 + \rho_2 + \mu)^2}\frac{\rho_2 + \mu}{\rho_3 + \mu} \end{array} \right] \end{array} \right\} \qquad (41)$$

For R to be a decreasing function of administration rate, i.e., $\frac{\partial \Re}{\partial \omega_1} < 0$, the given condition depicted in (42) needs to be met:

$$\frac{\mu\varepsilon_{V_3} + \rho_3\varepsilon_3}{\rho_3 + \mu} < \frac{\mu\varepsilon_{V_2} + \rho_2\varepsilon_2}{\rho_2 + \mu} \qquad (42)$$

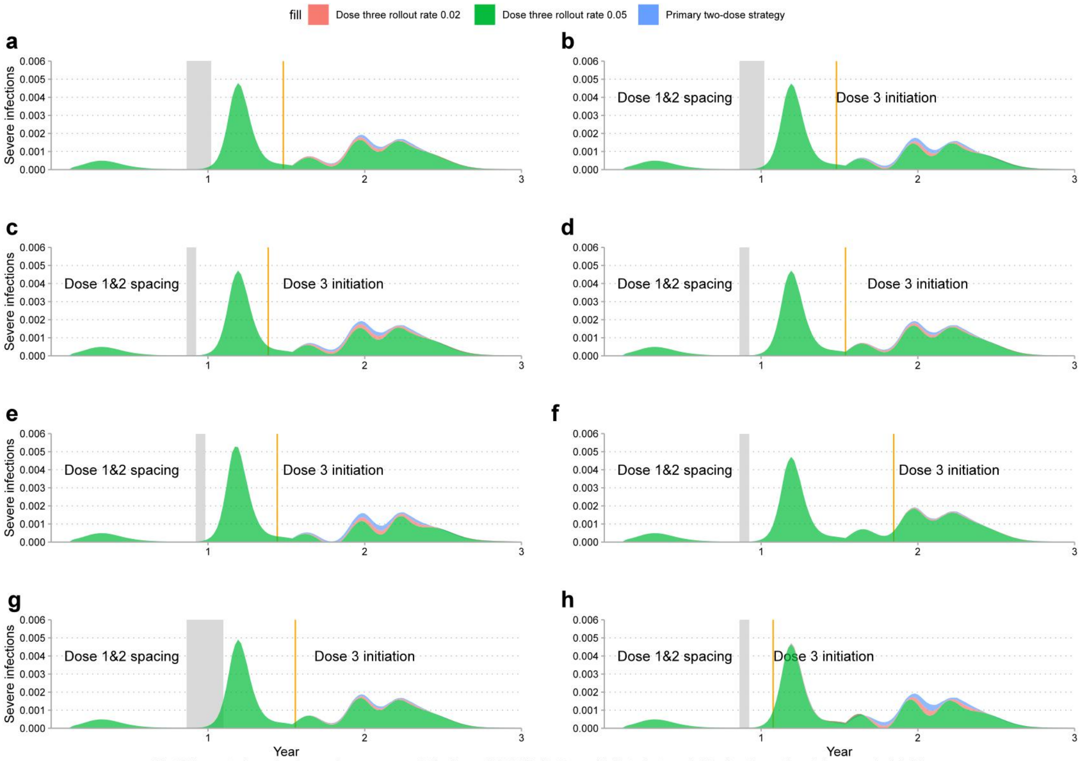


Fig.S2 Impact of vaccination regime on severe infections of COVID-19. Dose 1 initiated at week 45 after the outbreak in scenario (a)~(d) and (f)~(h). Initiation of dose 1 in scenario (e) delayed to week 47.The immunity conferred by dose 1 wanes to susceptibility after 6.5 weeks in all scenarios.
(a) Dose 1&2 spacing 8 weeks and 2&3 spacing 24 weeks, immunity conferred by dose 2&3 wanes to susceptibility after 26 weeks;
(b) Dose 1&2 spacing 8 weeks and 2&3 spacing 24 weeks, immunity conferred by dose 2&3 wanes to susceptibility after 52 and 26 weeks respectively;
(c) Dose 1&2 spacing 3 weeks and 2&3 spacing 24 weeks, immunity conferred by dose 2&3 wanes to susceptibility after 26 weeks;
(d) Dose 1&2 spacing 3 weeks and 2&3 spacing 32 weeks, immunity conferred by dose 2&3 wanes to susceptibility after 26 weeks;
(e) Dose 1&2 spacing 3 weeks and 2&3 spacing 24 weeks, immunity conferred by dose 2&3 wanes to susceptibility after 26 weeks;
(f) Dose 1&2 spacing 3 weeks and 2&3 spacing 48 weeks, immunity conferred by dose 2&3 wanes to susceptibility after 26 weeks;
(g) Dose 1&2 spacing 12 weeks and 2&3 spacing 24 weeks, immunity conferred by dose 2&3 wanes to susceptibility after 26 weeks;
(h) Dose 1&2 spacing 3 weeks and 2&3 spacing 8 weeks, immunity conferred by dose 2&3 wanes to susceptibility after 26 weeks.

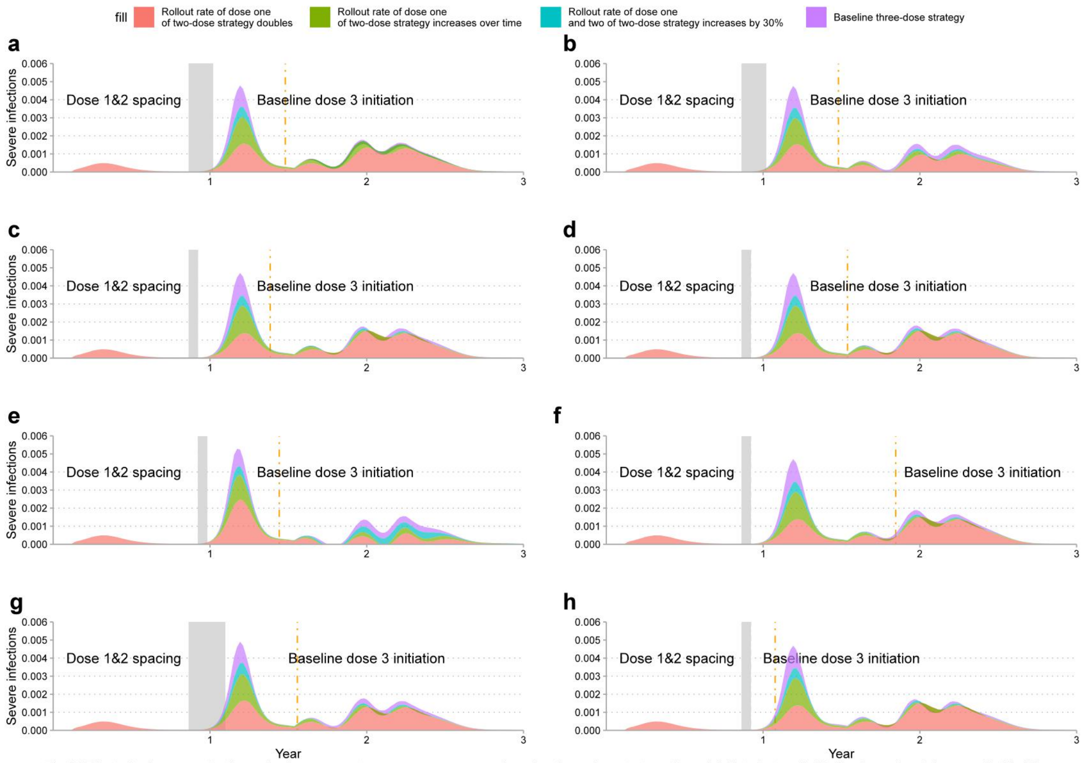


Fig.S3 Effect of primary vaccination series improvement on severe cases versus baseline three-dose strategy. Dose 1 initiated at week 45 after the outbreak in scenario (a)~(d) and (f)~(h). Initiation of dose 1 in scenario (e) delayed to week 47.The immunity conferred by dose 1 wanes to susceptibility after 6.5 weeks in all scenarios.
(a) Dose 1&2 spacing 8 weeks and 2&3 spacing 24 weeks, immunity conferred by dose 2&3 wanes to susceptibility after 26 weeks;
(b) Dose 1&2 spacing 8 weeks and 2&3 spacing 24 weeks, immunity conferred by dose 2&3 wanes to susceptibility after 52 and 26 weeks respectively;
(c) Dose 1&2 spacing 3 weeks and 2&3 spacing 24 weeks, immunity conferred by dose 2&3 wanes to susceptibility after 26 weeks;
(d) Dose 1&2 spacing 3 weeks and 2&3 spacing 32 weeks, immunity conferred by dose 2&3 wanes to susceptibility after 26 weeks;
(e) Dose 1&2 spacing 3 weeks and 2&3 spacing 24 weeks, immunity conferred by dose 2&3 wanes to susceptibility after 26 weeks;
(f) Dose 1&2 spacing 3 weeks and 2&3 spacing 48 weeks, immunity conferred by dose 2&3 wanes to susceptibility after 26 weeks;
(g) Dose 1&2 spacing 12 weeks and 2&3 spacing 24 weeks, immunity conferred by dose 2&3 wanes to susceptibility after 26 weeks;
(h) Dose 1&2 spacing 3 weeks and 2&3 spacing 8 weeks, immunity conferred by dose 2&3 wanes to susceptibility after 26 weeks.

**Table S1 Simulation parameters for Fig. 2**

| Name | Description | Baseline Value | References |
|---|---|---|---|
| $\alpha$ | relative infectiousness of $I_S$ | 1 | [17][45][46] |
| $\beta$ | transmission rate | $R*\gamma$ | [17] |
| $\alpha_V$ | relative infectiousness of $I_V$ | 1 | Assumed |
| $\alpha_1$ | relative infectiousness of $I_{S_1}$ | 1 | Assumed |
| $\alpha_2$ | relative infectiousness of $I_{S_2}$ | 1 | Assumed |
| $\alpha_3$ | relative infectiousness of $I_{S_3}$ | 1 | Assumed |
| $\varepsilon_1$ | susceptibility after the waning of dose 1 immunity | 0.5 | Assumed |
| $\varepsilon_2$ | susceptibility after the waning of dose 2 immunity | 0.5 | Assumed |
| $\varepsilon_3$ | susceptibility after the waning of dose 3 immunity | 0.1 | Assumed |
| $\varepsilon_{V_1}$ | infection rate after vaccination with dose 1 | 0.1 | Assumed |
| $\varepsilon_{V_2}$ | infection rate after vaccination with dose 2 | 0.05 | Assumed |
| $\varepsilon_{V_3}$ | infection rate after vaccination with dose 3 | 0.05 | Assumed |
| $\frac{1}{\rho_1}$ | the inverse of dose 1 immunity waning rate | 6.5 weeks | Assumed |
| $\frac{1}{\rho_2}$ | the inverse of dose 2 immunity waning rate | {26,52 }[weeks] Varies in the model. | Assumed |

| | | | |
|---|---|---|---|
| $\frac{1}{\rho_3}$ | the inverse of dose 3 immunity waning rate | 26 weeks | Assumed |
| $\mu$ | birth rate or death rate of population | 0.02 per week | [17][45][46] |
| $\gamma$ | the recovery rate of COVID-19 positive patients | 1.4 | [17][45][46] |
| $\nu$ | vaccination rate of dose 1 | 0.01 per week | [17][45][46] |
| $\omega$ | vaccination rate of dose 2 | 0.05 per week | Assumed |
| $\omega_1$ | vaccination rate of dose 3 | {0,0.02,0.05} [per week].<br>Varies in the model. | Assumed |
| $\delta$ | waning rate to secondary susceptibility | 1/3 | Assumed |
| $\{S_{vax_1}, S_{vax_2}, S_{vax_3}\}$ | initiation of dose 1&2&3 | {45,53,77}[week]<br>{45,48,72}<br>{45,48,80}<br>{45,48,96}<br>{47,50,74}<br>Varies in the model. | Assumed |
| $\mathbf{S}_P$ | initial size of the full susceptible population | 1- $I_0$ | [17] |
| $N$ | size of population | 1 | [17][45][46] |
| $I_0$ | initial size of infection | 1e-9 | [17] |
| $R$ | reproduction number | 2.3 | [17] |
| $c$ | partially susceptible individuals vaccinated and immunity equivalent to dose 1 | 0.01 | Assumed |
| $d$ | partially susceptible individuals vaccinated and immunity equivalent to dose 1 | 0.01 | Assumed |

### Table S2 Simulation parameters for Fig. 3

| Name | Description | Baseline Value | References |
|---|---|---|---|
| $\nu$ | vaccination rate of dose 1 | 0.01 per week | Baseline three-dose strategy |
| $\omega$ | vaccination rate of dose 2 | 0.05 per week | |
| $\omega_1$ | vaccination rate of dose 3 | 0.02 [per week]. | |
| $\nu$ | vaccination rate of dose 1 | 0.013 per week | Efficiency-enhanced primary vaccination case 1: rate of dose 1&2 increases by 30% |
| $\omega$ | vaccination rate of dose 2 | 0.05*1.3 per week | |
| $\omega_1$ | vaccination rate of dose 3 | 0 [per week]. | |
| $\nu$ | vaccination rate of dose 1 | $\nu \cdot 1.5^{\frac{t}{t+1}}$ | Efficiency-enhanced primary vaccination case 2: rate of dose 1 increases over time |
| $\omega$ | vaccination rate of dose 2 | 0.05 per week | |
| $\omega_1$ | vaccination rate of dose 3 | 0 [per week]. | |
| $\nu$ | vaccination rate of dose 1 | 0.02per week | Efficiency-enhanced primary vaccination case 3: rate of dose 1 doubles |
| $\omega$ | vaccination rate of dose 2 | 0.05 per week | |
| $\omega_1$ | vaccination rate of dose 3 | 0 [per week]. | |

### Table S3 Simulation parameters for Fig. 4

| **Name** | **(A) Severe Variant** | **(B)** | **(C)** | **(D)** |
|---|---|---|---|---|
| $\beta$ | 3.4 | 3.4 | 3.4 | 3.4 |
| $\varepsilon_1$ | 0.6 | 0.5 | 0.4 | 0.4 |
| $\varepsilon_2$ | 0.5 | 0.2 | 0.2 | 0.2 |
| $\varepsilon_3$ | 0.5 | 0.1 | 0.1 | 0.1 |
| $\varepsilon_{V_1}$ | 0.8 | 0.4 | 0.3 | 0.2 |
| $\varepsilon_{V_2}$ | 0.7 | 0.1 | 0.05 | 0.03 |
| $\varepsilon_{V_3}$ | 0.6 | 0.08 | 0.05 | 0.02 |
| $\frac{1}{\rho_1}$ | {5.2,13,26} weeks | {5.2,13,26} weeks | {5.2,13,26} weeks | {5.2,13,26} weeks |
| $\frac{1}{\rho_2}$ | 26 weeks | 26 weeks | 52 weeks | 78 weeks |
| $\frac{1}{\rho_3}$ | 26 weeks | 26 weeks | 52 weeks | 78 weeks |
| $\omega_1$ | 5e(-7)(Close to boundary). | 5e(-7)(Close to boundary). | 5e(-7)(Close to boundary). | 5e(-7)(Close to boundary). |

### Table S4 Simulation parameters for Fig. 5

| Name | (A) | (B) | (C) |
|---|---|---|---|
| $\beta$ | 3.4 | 3.4 | 3.4 |
| $\varepsilon_1$ | 0.5 | 0.5 | 0.5 |
| $\varepsilon_2$ | 0.3 | 0.3 | 0.3 |
| $\varepsilon_3$ | 0.1 | 0.08 | 0.05 |
| $\varepsilon_{V_1}$ | 0.1 | 0.1 | 0.1 |
| $\varepsilon_{V_2}$ | 0.05 | 0.05 | 0.05 |
| $\varepsilon_{V_3}$ | 0.03 | 0.02 | 0.01 |
| $\frac{1}{\rho_1}$ | 26 weeks | 26 weeks | 26 weeks |
| $\frac{1}{\rho_2}$ | {13,26,52} weeks | {13,26,52} weeks | {13,26,52} weeks |
| $\frac{1}{\rho_3}$ | 52 weeks | 52 weeks | 52 weeks |
| $\nu$ | 0.01 per week | 0.015 per week | 0.02 per week |

### Table S5 Simulation parameters for Fig. S2 and Fig. S3

| Name | Description | Value | References |
|---|---|---|---|
| $\lambda_P$ | Likelihood of being severe infection at $I_P$ | 0.14 | [17][45][46] |
| $\lambda_S$ | Likelihood of being severe infection at $I_S$ | 0.07 | [17][45][46] |
| $\lambda_V$ | Likelihood of being severe infection at $I_V$ | 0.1 | [17][45][46] |
| $\lambda_1$ | Likelihood of being severe infection at $I_V$ | 0.1 | [17][45][46] |
| $\lambda_2$ | Likelihood of being severe infection at $I_V$ | 0.1 | [17][45][46] |
| $\lambda_3$ | Likelihood of being severe infection at $I_V$ | 0.1 | Assumed |

### References


1. World Health Organization. WHO Coronavirus (COVID-19) Dashboard. https://covid19.who.int (2022).
2. Wouters, O. J. et al. Challenges in ensuring global access to COVID-19 vaccines: production, affordability, allocation, and deployment. Lancet 397,1023–34(2021).
3. World Health Organization. WHO COVID-19 vaccines https://www.who.int/emergencies/diseases/novel-coronavirus-2019/covid-19-vaccines (2022).
4. Chung. J. Y. et al. COVID-19 vaccines: The status and perspectives in delivery points of view. Adv. Drug. Deliv. Rev. 170, 1–25 (2021).
5. MEO, S. A. et al. COVID-19 vaccines: comparison of biological, pharmacological characteristics and adverse effects of Pfizer/BioNTech and Moderna Vaccines. Eur. Rev. Med. Pharmacol. Sci. 25, 1663-1669 (2021).
6. World Health Organization. WHO COVID-19 advice for the public. https://www.who.int/emergencies/diseases/novel-coronavirus-2019/covid-19-vaccines/advice (2022).
7. Mathieu, E. et al. A global database of COVID-19 vaccinations. Nat. Hum. Behav. 5,947-953 (2021).
8. Our World in Data.Statistics and Research Coronavirus (COVID-19) Vaccinations.https://ourworldindata.org/covid-vaccinations (Accessed on January 5, 2022).
9. Harrison, E. A. et al. Vaccine confidence in the time of COVID-19. Eur. J. Epidemiol. 35, 325-330 (2020) .
10. Jecker, N. S. et al. Three for me and none for you? An ethical argument for delaying COVID-19 boosters. J. Med. Ethics. ,10.1136/medethics-2021-107824 (2021).
11. World Health Organization. Interim statement on booster doses for COVID-19 vaccination. https://www.who.int/news/item/22-12-2021-interim-statement-on-booster-doses-for-covid-19-vaccination---update-22-december-2021 (2022).
12. Polack, F. P. et al. Safety and Efficacy of the BNT162b2 mRNA Covid-19 Vaccine. N. Engl. J. Med. 383, 2603-15 (2020).
13. Vasileiou, E. et al. Interim findings from first-dose mass COVID-19 vaccination roll-out and COVID-19 hospital admissions in Scotland: a national prospective cohort study. Lancet 397, 1646–57(2021).
14. Lamb, Y.N. et al. BNT162b2 mRNA COVID-19 Vaccine: First Approval. Drugs 81, 495 - 501 (2021) .
15. Sadoff, J. Safety and Efficacy of Single-Dose Ad26.COV2.S Vaccine against Covid-19. N. Engl. J. Med. 384,2187-201 (2021).
